# Epidemiological impact and cost-effectiveness analysis of COVID-19 vaccination in Kenya

**DOI:** 10.1101/2022.04.21.22274150

**Authors:** Stacey Orangi, John Ojal, Samuel P. C. Brand, Cameline Orlendo, Angela Kairu, Rabia Aziza, Morris Ogero, Ambrose Agweyu, George M Warimwe, Sophie Uyoga, Edward Otieno, Lynette I Ochola-Oyier, Charles N Agoti, Kadondi Kasera, Patrick Amoth, Mercy Mwangangi, Rashid Aman, Wangari Ng’ang’a, Ifedayo M O Adetifa, J Anthony G Scott, Philip Bejon, Matt. J. Keeling, Stefan Flasche, D. James. Nokes, Edwine Barasa

## Abstract

**Background:** Few studies have assessed the benefits of COVID-19 vaccines in settings where most of the population had been exposed to SARS-CoV-2 infection.

**Methods:** We conducted a cost-effectiveness analysis of COVID-19 vaccine in Kenya from a societal perspective over a 1.5-year time frame. An age-structured transmission model assumed at least 80% of the population to have prior natural immunity when an immune escape variant was introduced. We examine the effect of slow (18 months) or rapid (6 months) vaccine roll-out with vaccine coverage of 30%, 50% or 70% of the adult (> 18 years) population prioritizing roll-out in over 50-year olds (80% uptake in all scenarios). Cost data were obtained from primary analyses. We assumed vaccine procurement at $7 per dose and vaccine delivery costs of $3.90-$6.11 per dose. The cost-effectiveness threshold was USD 919.

**Findings:** Slow roll-out at 30% coverage largely targets over 50-year-olds and resulted in 54% fewer deaths (8,132(7,914 to 8,373)) than no vaccination and was cost-saving (ICER=US$-1,343 (-1,345 to - 1,341) per DALY averted). Increasing coverage to 50% and 70%, further reduced deaths by 12% (810 (757 to 872) and 5% (282 (251 to 317) but was not cost-effective, using Kenya’s cost-effectiveness threshold ($ 919.11). Rapid roll-out with 30% coverage averted 63% more deaths and was more cost-saving (ICER=$-1,607 (-1,609 to -1,604) per DALY averted) compared to slow roll-out at the same coverage level, but 50% and 70% coverage scenarios were not cost-effective.

**Interpretation:** With prior exposure partially protecting much of the Kenyan population, vaccination of young adults may no longer be cost-effective.

**KEY QUESTIONS:** *What is already known?:*

- The COVID-19 pandemic has led to a substantial number of cases and deaths in low-and middle-income countries.
- COVID-19 vaccines are considered the main strategy of curtailing the pandemic. However, many African nations are still at the early phase of vaccination.
- Evidence on the cost-effectiveness of COVID-19 vaccines are useful in estimating value for money and illustrate opportunity costs. However, there is a need to balance these economic outcomes against the potential impact of vaccination.

*What are the new findings?:*

- In Kenya, a targeted vaccination strategy that prioritizes those of an older age and is deployed at a rapid rollout speed achieves greater marginal health impacts and is better value for money.
- Given the existing high-level population protection to COVID-19 due to prior exposure, vaccination of younger adults is less cost-effective in Kenya.

*What do the new findings imply?:*

- Rapid deployment of vaccines during a pandemic averts more cases, hospitalisations, and deaths and is more cost-effective.
- Against a context of constrained fiscal space for health, it is likely more prudent for Kenya to target those at severe risk of disease and possibly other vulnerable populations rather than to the whole population.

## INTRODUCTION

As of early April 2022, Kenya has experienced five distinct waves of the COVID-19 pandemic with more than 320,000 reported cases and 5,600 deaths.[1] While at the global level vaccines to prevent severe disease from SARS-CoV-2 are the main strategy for curtailing the pandemic burden on health,[2] most African nations are still at a very early phase of vaccine roll-out, particularly in tropical sub-Saharan Africa, with most countries at less than 10% of the adult population fully vaccinated.[1] However, in contrast to other part of the world where low vaccine coverage in high risk groups has led to high mortality even from the omicron variant,[3] in Kenya cross-sectional serological surveys of anti-SARS-CoV-2 spike protein antibodies together with transmission dynamic model forecasts indicate that about 80% of the population have been exposed to the virus at least once and thus generated considerable immunity[4] with similar estimates in the region.[5] This raises the question what additional benefit can vaccination still have in mitigating future disease burden from COVID-19?

The Kenyan government is pursuing a phased COVID-19 vaccination strategy that aims to follow a risk-prioritization matrix leading sequentially to the vaccination of all adults by December 2022.[6] The prioritized population are an estimated 30% of the adult population and include health and other essential workers, individuals at high risk of severe disease (those above 58 years, and those above 18 years with co-morbidities), and individuals at high risk of infection (individuals in congregate settings, and those working in hospitality and transport sectors).[6] Vaccine roll-out commenced in early March 2021. As of early April, 2022 more than 17.7 million doses had been administered with 30% of Kenya’s adult population above the age of 18 years being fully vaccinated.[7] The initial procurement comprised of the Oxford/Astra Zeneca vaccine mainly sourced through the COVID-19 Vaccines Global Access Facility (COVAX) mechanism and bilateral negotiations, evolving more recently to a multi-vaccine type deployment through additional sources including the African Union’s (AU) African Vaccine Acquisition Task Team (AVATT) mechanism.[6]

Economic evaluations are useful in providing evidence of the value for money for different health interventions and illustrates the opportunity costs of the interventions in a setting with many competing priorities. However, there is a need to balance these economic outcomes against the potential impact of the interventions. Therefore, this study evaluates the potential epidemiological impact and cost-effectiveness of different vaccine roll-out scenarios in a Kenyan population that has already acquired a high-level immunity due to prior infections. The study employs a partially retrospective perspective with vaccination scenarios beginning September 2021 and with an immune escape variant striking in November 2021.

## METHODS

### Study design

This study is an impact and cost-effectiveness analysis of COVID-19 vaccine roll-out strategies that uses cost estimates from primary costing studies and vaccine effectiveness measures from an age structured transmission model. The costs and effects are estimated from a societal perspective for a period of 1.5 years (1^st^ September 2021 to 28^th^ February 2023) starting at the peak of the Kenyan delta wave and simulating the emergence of a partial immune escape variant (omicron-like) from November 2021.

### Intervention comparators

#### i. Primary analysis

We carry out an incremental analysis of four vaccination coverage scenarios deployed over an 18-month period (non-rapid deployment), starting at 0% coverage in September 2021 (Table 1): No vaccination (0% coverage), or 30%, 50% and 70% coverage of the population older than 18 years with prioritization of those aged 50 years and above (until 80% of those >50y old are fully vaccinated), then the remaining doses given to those 18-49y old).

**Table 1:** Intervention comparators and number vaccinated within 1.5 years’ time horizon

| Vaccination strategy | Number of >50y olds who were vaccinated (proportion of those>50y) | Number of 18-49y olds who were vaccinated (proportion of those 18-49y) |
| --- | --- | --- |
| No vaccination |  |  |
| 30% adult coverage strategy | 4,133,775 (80%) | 3,186,225 (19%) |
| 50% adult coverage strategy | 4,133,775 (80%) | 8,366,225 (41%) |
| 70% adult coverage strategy | 4,133,775 (80%) | 13,366,225 (65%) |

#### ii. Secondary analysis

We consider a secondary analysis that assesses the same scenarios under the primary analysis but with rapid vaccine deployment in which the targeted vaccine coverage is attained within 6 months of starting vaccination.

An assumption was made that all the vaccination coverage scenarios and deployment strategies were implemented alongside a low intensity mix of non-pharmaceutical interventions (NPI). The low intensity NPI is matched with how government progressively lifted or modified the restrictions and refers to re-opening of international borders, relaxed curfew, controlled public gatherings, controlled re-opening of restaurants and bars, controlled re-opening of schools, ban lift on mandatory use of masks, and adherence to hand hygiene from November 2020 to the time of writing this manuscript.

### Transmission modelling and parameter inference

We extended a dynamic SARS-CoV-2 transmission model previously designed to estimate population level immunity from natural infection in Kenya by fitting to case notification and serological data[3] to include additional age structure and vaccination status. In common with other approaches to modelling SARS-CoV-2 transmission,[8,9] we assume that the rate of new infections depends on: (i) age and setting-specific contact rates within the population, (ii) frequency of Alpha, Beta and Delta variants of SARS-CoV-2 among the infected sub-population, (iii) the first and second dose vaccine protection against infection in each age group which were assumed to wane over time, and, (iv) prior primary infections. The probability of being infected with SARS-CoV-2 per infectious contact, and the chance of developing symptoms upon infection, increased substantially with age (see *supplementary information* for details of the transmission model).

The goal of the transmission model is to project the health gains of the vaccine deployment strategies described above in comparison with the no vaccine scenario. This requires estimation of parameters pertaining to the risk of transmission, and, of risk factors associated with infection given age, and the infecting variant of SARS-CoV-2 in the Kenyan setting. These parameters were inferred by fitting the model to the following Kenyan epidemiological data (see SI for inference methods):

- Daily reported numbers of positive and negative PCR tests from the Kenyan Ministry of Health COVID-19 linelist (between 1^st^ January 2021 and 1^st^ November 2021).
- Cross-sectional serological surveys of (a) donor samples from the Kenyan National Blood and Transfusion Service (KNBTS)[10] and (b) demographic surveillance systems (between 1^st^ January 2021 and 27^th^ May 2021).[11]

We used a Bayesian hierarchical inference approach aimed at allowing inference on reporting fraction in counties with higher numbers of serological tests to influence inference of reporting fraction in counties with lower numbers of serological tests (*see supplementary information* for details on underlying data for age-specific effects and details on inference methodology).

### Infection outcome modelling and risk factor inference

Bayesian inference of transmission model parameters generated a posterior predictive distribution for the number of SARS-CoV-2 infections in Kenya broken down by day, county, age of infected, infecting variant of SARS-CoV-2, and, whether it was a primary infection event or a re-infection event. We categorized the outcome of each infection as being either deadly, critical (requiring treatment in an intensive care unit (ICU)), severe (requiring in-patient hospitalization in a general ward), mild or asymptomatic. Severe and critical infections were assumed to cause admission to a health facility’s general ward or ICU for an average of 12 days post-infection. Severe infection was assumed to lead to an average 7 day stay in a general COVID ward before discharge. Critical infection leads to an average 7 day stay in ICU,[12] before transfer to a general COVID ward for a further average 7 day stay before discharge (see *supplementary information* for details on hospital durations of stay).

Risk factors for infection outcome were inferred using reported Kenya outcome data:

- Daily reported numbers occupying general health facilities with COVID-19 as the diagnosed cause (1^st^ March 2021 – 1^st^ November 2021).
- Daily reported numbers occupying ICUs with COVID-19 as the diagnosed cause (1^st^ March 2021 – 1^st^ November 2021).
- Daily reported incidence of death with COVID-19 as the diagnosed cause (1^st^ January 2021 – 1^st^ November 2021).

### Vaccination rollout modelling

We used the fitted model to predict the course of the pandemic from 1^st^ September 2021 (historically this was past the peak of the fourth wave of cases in Kenya) to 30 June 2023 and the impact of vaccination on, severe and critical disease, and deaths. We distribute the total number of doses planned under each vaccination scenario to the 47 counties proportionally according to population size above the age of 18 years.[13] We assume that the number of doses given per day will be the same during the study period. Doses will be offered to adults older than 50 years first, until take up of available vaccines dropped off, which we assumed would occur once 80% of over 50s had taken up both doses. The remaining doses will subsequently be randomly allocated to all 18-to 50-year-olds. Within the model, individuals are either unvaccinated, partially vaccinated (14 days after receipt of the first dose), fully vaccinated (14 days after receipt of the second dose) or have waned vaccine effectiveness. We assumed vaccine effectiveness against death (delta variant) to range from 90% to 95% after the first dose and 95% to 99% after the second dose.[14] Vaccine effectiveness against severe or critical disease (delta variant) of 80% to 90% and 95% to 99% after the first and second dose respectively.[14] The vaccine effectiveness against acquisition of infection per infectious contact (delta variant) was 55% to 65% and 65% to 80% after the first and second dose respectively.[14] We assumed an effectiveness of 0% to 35% and 0% to 69% against onward transmission, per infection (delta variant), after either the first or second dose.[15] We assume that immunity due to either past infection of vaccination eventually wanes to 70% protection against disease and 0% protection against infection, with a mean time to complete waning of 460 days after the second dose of vaccine and 5 years following natural infection.[16] Furthermore, we assume that protection due to prior infection combined constructively with vaccination; that is that people who had previously had a natural infection episode of SARS-CoV-2 were further protected from reinfection by vaccination (see *supporting information*).

### Immune escape variant

The scenarios investigated in this paper involve the rapid spread of a new variant of SARS-CoV-2 that, due to evolutionary adaptation, partially avoids protection from infection due to prior naturally acquired immunity and/or vaccination. Concretely, we assume that the immune escape variant enters Kenya in early November 2021 and rapidly dominates transmission by 15^th^ November 2021. Compared to homologous protection against reinfection with the Delta variant, the protection afforded by prior infection and/or vaccination against acquiring the novel immune escape variant is assumed to be decreased by 50%, with all epidemiological rates increased such that the mean generation time of transmission is reduced by 30% compared to the transmission of the Delta variant. However, we also assume that the fundamental reproductive number and risk factors for severe, critical and deadly outcomes are unchanged compared to Delta. (see *supplementary information* for the details of how a 50% decrease in protection from infection was implemented).

### Cost estimates

The cost estimates used in this study were derived using a hybrid method that involved both an ingredients approach (bottom-up) and a top-down approach.[17,18] The analysis used economic costs, which reflect the opportunity cost and incorporated both recurrent and capital costs. Capital costs were annuitized using a discount rate of 3% over their useful life. Costs incurred in other years were adjusted for inflation using the Gross domestic product (GDP) deflator and reported in 2021 United States Dollars (USD). Key model cost input parameters are shown in Table 2 and the three main cost components are described below. The costs of NPIs were excluded as all vaccination strategies employed the same NPI regime (low NPI intensity) and would therefore not change the reported incremental cost-effectiveness ratios (ICERs).

**Table 2:** Key analysis parameters

| Parameter | Values (Lb; Ub) | Source |
| --- | --- | --- |
| **Cost-effectiveness parameters** |  |  |
| Treatment costs (2021 US\$) | | |
| Per day, per patient unit cost of management of asymptomatic COVID-19 | \$19.75*testing rate | [20] |
| Per day, per patient unit cost of management of mild to moderate COVID-19 | \$19.75*testing rate | [20] |
| Per day, per patient unit cost of management of severe COVID-19 | \$129.45 | [20] |
| Per day, per patient unit cost of management of critical COVID-19 | \$623.14 | [20] |
| Testing rate |  |  |
| *Testing rate in the population | 0.52% | Proportion of reported to modelled cases |
| Vaccination costs (2021 US\$) | | |
| Vaccine procurement costs per dose | \$8.67 (Base cost: \$7 and including importation costs) | [19] |
| Supplies procurement costs per dose | \$0.08 | [19] |
| Vaccine delivery cost per dose (no vaccination) | \$0 | [19] |
| Vaccine delivery cost per dose (30% coverage) | \$6.11 | [19] |
| Vaccine delivery cost per dose (50% coverage) | \$4.16 | [19] |
| Vaccine delivery cost per dose (70% coverage) | \$3.90 | [19] |
| Duration of disease and length of hospitalization |  |  |
| Length of hospitalization for severe episode | 7 days (4-11) | Assumption |
| Length of ICU stay for critical episode | 7 days (4-11) | [12] |
| Duration of asymptomatic disease | 7 days | Assumption |
| Duration of mild to moderate disease | 7 days | Assumption |
| Duration of severe disease | 12 days | [24] |
| Duration of critical disease | 20 days | [24] |
| DALYs |  |  |
| Disability weight for asymptomatic episode | 0 |  |
| Disability weight for mild/moderate episode | 0.051 (0.032; 0.074) | [25] |
| Disability weight for severe episode | 0.133 (0.088; 0.191) | [25] |
| Disability weight for critical episode | 0.655 (0.579; 0.727) | [26] |
| *Average age at death |  | [13] |
| 0-19 years | 9.27years |  |
| 20-49 years | 31.75 years |  |
| 50-59 years<br>60-69 years<br>70-79 years<br>80+ years | 54.10 years<br>63.85 years<br>73.41 years<br>86.00 years |  |
| Life expectancy<br>0-19 years<br>20-49 years<br>50-59 years<br>60-69 years<br>70-79 years<br>80+ years | 64.10 years<br>40.94 years<br>24.71 years<br>17.64 years<br>11.41 years<br>4.84 years | [29] |
| Cost-effectiveness threshold per DALY averted | 0.5*GDP per capita.<br>[GDP per capita = \$1,838.21] | [27,28,30] |
| **Transmission dynamic model parameters** |  |  |
| Transmission dynamic model values | See Table S1 |  |

\*Testing rate: A proxy estimate is used that is calculated as a proportion of reported cases to modelled cases across all severity levels from Jan1 to Sep 19, 2021.
\*Average age at death is based on the weighted mean age across the different age groups

#### i. Vaccination costs

We included vaccine and related supplies costs, as well as vaccine delivery costs. Vaccine and related supply costs were the economic costs to purchase the vaccine and related supplies such as syringes and safety boxes through the COVAX facility. A base cost of $7 was used for vaccine procurement, which is the country’s procurement cost from the COVAX facility. Additionally, the freight costs, insurance costs, import declaration fees, clearance fees, and the railway development levy associated with the vaccines and its supplies were included. Vaccine and syringe wastage rates of 10% were assumed.[19] Vaccine delivery costs referred to costs associated with delivering COVID-19 immunizations to the adult population and were estimated across six components 1) vaccine supply chain 2) vaccine safety monitoring and adverse events following immunization management 3) training 4) advocacy, communication and social mobilization 5) data management, monitoring and supervision 6) vaccine administration. The resource used and costs were estimated through the analysis of programmatic budgets, and through key informant interviews. Details of the vaccine procurement and delivery cost analysis and results are reported elsewhere.[19] This analysis assumed equivalent vaccine delivery costs for both the rapid and non-rapid vaccination strategies.

#### ii. Treatment costs

The direct medical costs of COVID-19 treatment were sourced from a recently conducted study that examined the unit costs for COVID-19 case management in Kenya.[20] This costing analysis employed an ingredients-based approach to estimate health care costs across the disease severity categories; with the exclusion of adverse events costs.[20]

#### iii. Productivity losses

Productivity losses due to illness and mortality were estimated using a human capital approach.[21] The impact of COVID-19 on lost time through illness or morbidity was estimated by accounting for the average Kenyan’s productivity measure (GDP per capita) and duration of disease/duration of quarantine; the latter was used where duration of illness was less than the 14 day quarantine period in Kenya. For asymptomatic and mild disease, the testing rate was accounted for and an assumption was made that only those in the informal sector are likely not to be productive as they isolate. Further, the economic impact of COVID-19-related mortality was estimated by considering the years of life lost because of premature mortality and the average productivity measure. We did not account for productivity losses from long COVID, as the burden is poorly defined in our setting. (See equation (a) in the supplementary information)

### Disability-adjusted life-years

The outcome of the cost-effectiveness analysis was reported in terms of disability-adjusted life-years (DALYs); the sum of years of life lost (YLL) and years lost due to disability (YLD) [22]. (See equations (b), (c), and (d) in the supplementary information).

DALYs were calculated considering a discount rate of 3%, the Kenyan 2019 standardized life expectancies,[23] assumed duration of illness of 7 days for asymptomatic and mild disease and 12 and 20 days for severe and critical disease respectively,[24] as well as disability weights. COVID-19 is a novel disease, and its disability weights are currently not available. Therefore, for asymptomatic COVID-19 disease we assumed a disability weight of 0. For mild-to-moderate COVID-19 symptoms and severe disease, we used disability weights from the 2013 Global Burden of disease of 0.051 (0.032-0.074) and 0.133 (0.088-0.190) assigned to infectious disease with moderate acute episodes and severe episodes respectively.[25] For critical disease, we assume disability weights of 0.655 (0.579-0.727) assigned to intensive care unit admissions.[26] This analysis did not incorporate age-weighting in the DALYs. These input parameters are reported in Table 2.

The incremental cost-effectiveness ratio (ICER) was the measure of cost-effectiveness calculated as the net change in total costs and DALYs averted between comparators. The ICER was compared with the opportunity cost-based on Kenya’s cost-effectiveness threshold (USD 919.11).[27,28]

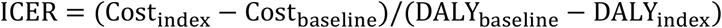

*Where*:

Cost_index_ =cost of strategy of interest

Cost_baseline_=cost of the next less effective strategy

DALY_index_= total DALYs under the strategy of interest.

DALY_baseline_=total DALYs under the next less effective strategy

ICERs are estimated within each of the two roll-out scenarios slow and rapid and are not comparable between the two vaccine-deployment cases except when the baseline is no vaccination.

### Sensitivity analysis for the model

Vaccine effectiveness against different epidemiological outcomes such as the acquisition of disease, onward transmission, severe disease and death does vary with age, duration between vaccination and testing of efficacy, variant of infection, and type of vaccine being used amongst other factors.[31–34] Therefore, to determine the robustness of the epidemiological model predictions to the vaccine effectiveness parameter values, we performed a sensitivity analysis across a range of values using a vaccine waning effectiveness model fitted to the UK Health Security Agency (UKHSA) COVID-19 data.[14,15]

A univariate sensitivity analysis was done on the economic model to determine the robustness of the unit cost estimates with variations in vaccine procurement costs (base cost of $3 and $10 used) and discounting rates of DALYs (rate of 0% used). Further, given the current evidence gap to confidently determine the magnitude of underreporting of COVID-19 deaths,[35] the baseline cost-effectiveness analysis assumed an under-reporting of hospitalization and deaths by a factor of 5 and a one-way sensitivity analysis was done by varying the under-reporting factor (1-4).

A probabilistic sensitivity analysis to explore the influence of some economic parameters on the ICERs was done using Sobol sampling and was based on the statistical distributions in Table S2. Sobol sequences belong to the family of quasi-random sequences which are designed to generate samples of multiple parameters as uniformly as possible over the multi-dimensional parameter space.[36] For the parameters used in the probabilistic sensitivity analysis, the statistical distributions were chosen to model the available prior knowledge represented by existing data, as reported in Table 2. For the cost estimates range, a 20% increase or decrease was assumed for the parameters.

### Stakeholder engagement

The results of the study have been disseminated to key policy makers and relevant stakeholders involved in COVID-19 vaccine deployment in Kenya.

## RESULTS

### Clinical Impacts of Vaccination Strategies and scenarios

The non-rapid deployment of vaccinating 30% of the adult population results in 10% (32 (24 to 38) per 100,000) fewer infections, 54% (8,132 (7,914 to 8,373) fewer deaths compared to no vaccination, and 978 (949 to 1,005) people would need to be vaccinated to prevent 1 death. An increase of vaccine coverage of the adult population to 50% results in a further 1% (4 (3 to 5) per 100,000) reduction in infections, a further 12% (810 (757 to 872) reduction in deaths, and 5,617 (5,218 to 6,011) more people would need to be vaccinated to prevent an additional death. Similarly, an increase of vaccine coverage to 70% leads to a 1% reduction in cases, a 5% reduction in deaths, and 17,730 (15,773 to 19,920) more people would need to be vaccinated to prevent an additional death compared to the 50% vaccination coverage.

In the rapid vaccine rollout strategy, the 30% vaccine coverage averts 12% of cases preventing an average of 39 (29 to 48) per 100,000 infections and 63% of deaths saving an average of 9,433 (9,197 to 9,711) lives compared to no vaccination. Therefore, 843 (819 to 864) people would need to be vaccinated to prevent a death. The 30% coverage under a rapid deployment averts more cases and saves more lives compared to a non-rapid rollout with the same level of coverage. See Table 3 and Figure 1.

**Table 3:**
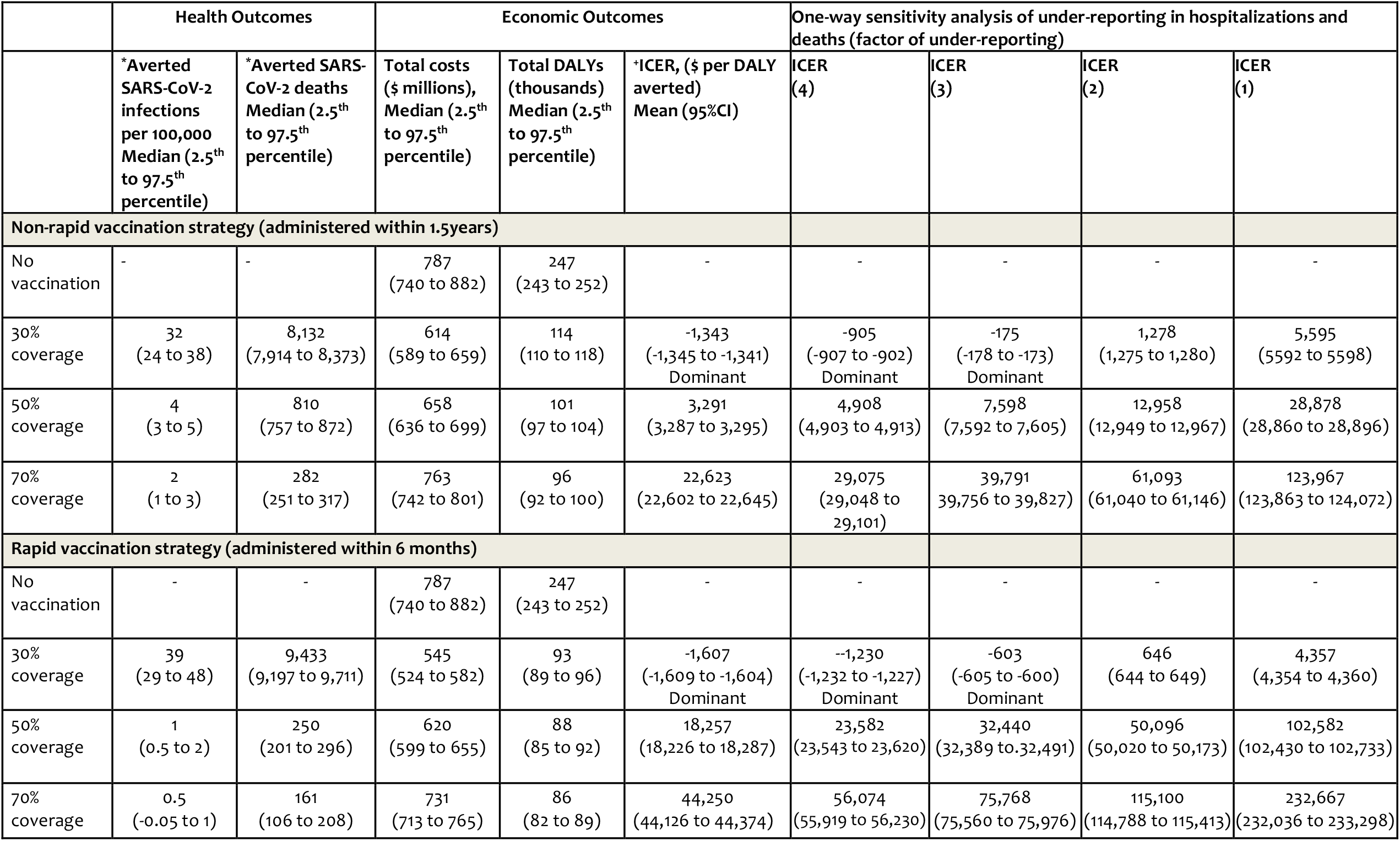
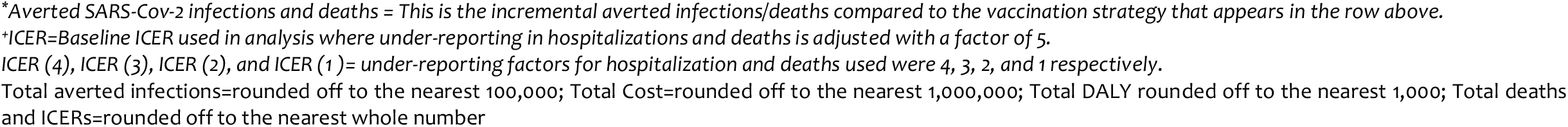
Projected clinical outcomes, costs, and the cost-effectiveness of different vaccination strategies in Kenya from a societal perspective

|  | Health Outcomes |  | Economic Outcomes |  |  | One-way sensitivity analysis of under-reporting in hospitalizations and deaths (factor of under-reporting) |  |  |  |
| --- | --- | --- | --- | --- | --- | --- | --- | --- | --- |
| | *Averted SARS-CoV-2 infections per 100,000 Median (2.5<sup>th</sup> to 97.5<sup>th</sup> percentile) | *Averted SARS-CoV-2 deaths Median (2.5<sup>th</sup> to 97.5<sup>th</sup> percentile) | Total costs (\$ millions), Median (2.5<sup>th</sup> to 97.5<sup>th</sup> percentile) | Total DALYs (thousands) Median (2.5<sup>th</sup> to 97.5<sup>th</sup> percentile) | *ICER, (\$ per DALY averted) Mean (95%CI) | ICER (4) | ICER (3) | ICER (2) | ICER (1) |
| **Non-rapid vaccination strategy (administered within 1.5years)** |  |  |  |  |  |  |  |  |  |
| No vaccination | - | - | 787 (740 to 882) | 247 (243 to 252) | - | - | - | - | - |
| 30% coverage | 32 (24 to 38) | 8,132 (7,914 to 8,373) | 614 (589 to 659) | 114 (110 to 118) | -1,343 (-1,345 to -1,341) Dominant | -905 (-907 to -902) Dominant | -175 (-178 to -173) Dominant | 1,278 (1,275 to 1,280) | 5,595 (5592 to 5598) |
| 50% coverage | 4 (3 to 5) | 810 (757 to 872) | 658 (636 to 699) | 101 (97 to 104) | 3,291 (3,287 to 3,295) | 4,908 (4,903 to 4,913) | 7,598 (7,592 to 7,605) | 12,958 (12,949 to 12,967) | 28,878 (28,860 to 28,896) |
| 70% coverage | 2 (1 to 3) | 282 (251 to 317) | 763 (742 to 801) | 96 (92 to 100) | 22,623 (22,602 to 22,645) | 29,075 (29,048 to 29,101) | 39,791 (39,756 to 39,827) | 61,093 (61,040 to 61,146) | 123,967 (123,863 to 124,072) |
| **Rapid vaccination strategy (administered within 6 months)** |  |  |  |  |  |  |  |  |  |
| No vaccination | - | - | 787 (740 to 882) | 247 (243 to 252) | - | - | - | - | - |
| 30% coverage | 39 (29 to 48) | 9,433 (9,197 to 9,711) | 545 (524 to 582) | 93 (89 to 96) | -1,607 (-1,609 to -1,604) Dominant | -1,230 (-1,232 to -1,227) Dominant | -603 (-605 to -600) Dominant | 646 (644 to 649) | 4,357 (4,354 to 4,360) |
| 50% coverage | 1 (0.5 to 2) | 250 (201 to 296) | 620 (599 to 655) | 88 (85 to 92) | 18,257 (18,226 to 18,287) | 23,582 (23,543 to 23,620) | 32,440 (32,389 to 32,491) | 50,096 (50,020 to 50,173) | 102,582 (102,430 to 102,733) |
| 70% coverage | 0.5 (-0.05 to 1) | 161 (106 to 208) | 731 (713 to 765) | 86 (82 to 89) | 44,250 (44,126 to 44,374) | 56,074 (55,919 to 56,230) | 75,768 (75,560 to 75,976) | 115,100 (114,788 to 115,413) | 232,667 (232,036 to 233,298) |

*\*Averted SARS-Cov-2 infections and deaths = This is the incremental averted infections/deaths compared to the vaccination strategy that appears in the row above.*
*\*ICER=Baseline ICER used in analysis where under-reporting in hospitalizations and deaths is adjusted with a factor of 5.*
*ICER (4), ICER (3), ICER (2), and ICER (1)= under-reporting factors for hospitalization and deaths used were 4, 3, 2, and 1 respectively.*
*Total averted infections=rounded off to the nearest 100,000; Total Cost=rounded off to the nearest 1,000,000; Total DALY rounded off to the nearest 1,000; Total deaths and ICERs=rounded off to the nearest whole number*

**Figure 1:**
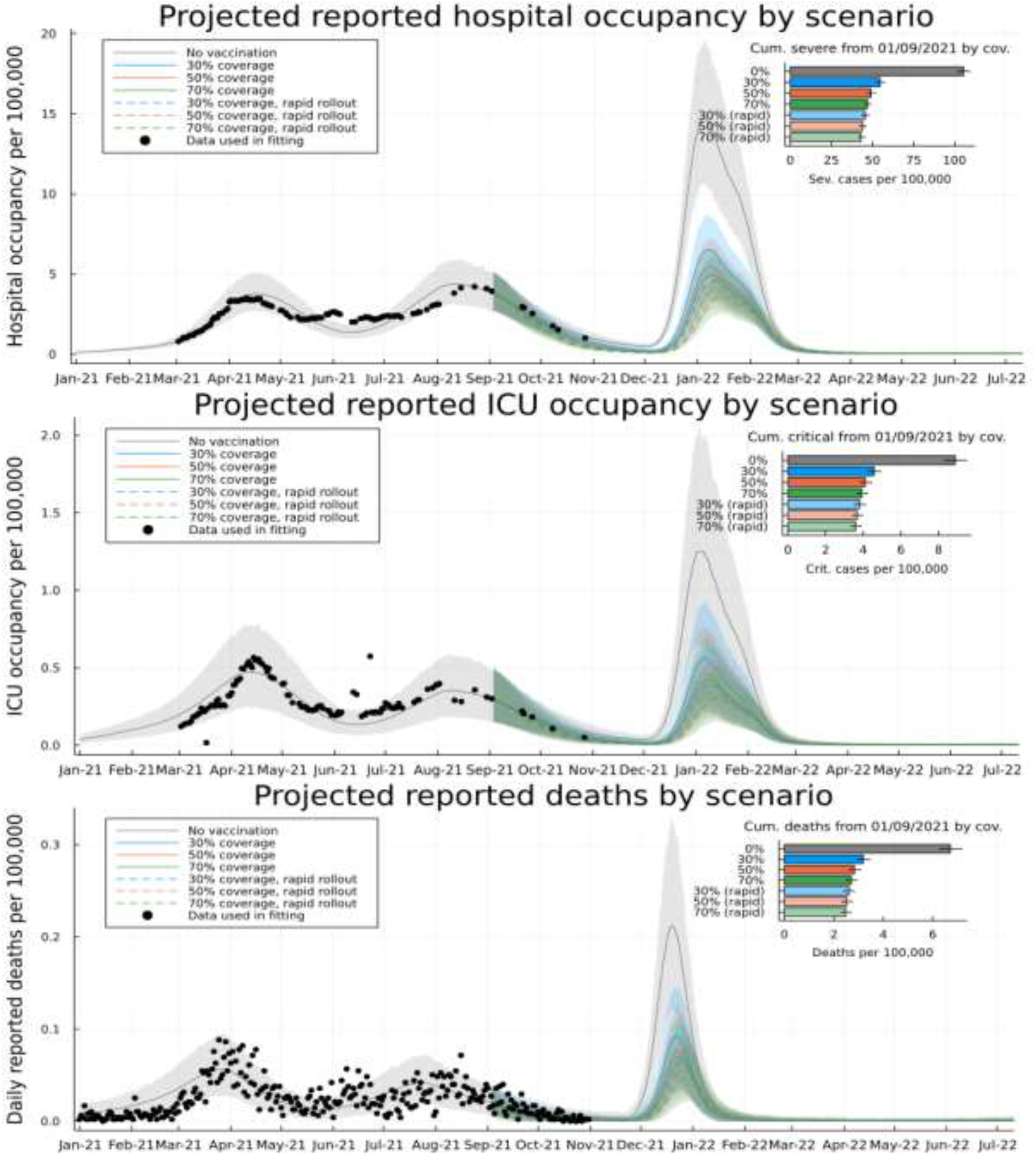
Model-based projections and vaccine scenarios: Model-based prediction intervals for daily occupancy of general wards in health facilities in Kenya (top), daily occupancy of intensive care units in Kenya (middle), and daily reported incidence of death with COVID in Kenya (bottom). All scatter points represented data used in inference of the infection outcome model. Grey curves are the posterior mean model prediction (background shading 95% CIs) with no vaccinations. Colored curves represent a target of 30% (blue), 50% (red) and 70% (green) of over 18 year old population in Kenya over 18 months (solid) or 6 months (dashed). Insets: Projections of cumulative number of severe (top), critical (middle) and deadly (bottom) cases after 1st September 2021 under each vaccine target scenario

### Cost-Effectiveness of Vaccination Strategies

Table 3 shows the total costs, DALYs and ICERs of the vaccination scenarios considered in the analysis from a societal perspective. Under the non-rapid vaccination scenario, vaccinating 30% of the adult population is cost-saving (ICER=$-1,343 (-1,345 to -1,341) per DALY averted) and hence highly cost-effective. Increasing vaccine coverage to 50% of the adult population was not cost-effective (ICER=$3,291 (3,287 to 3,295) per DALY averted) compared to 30% coverage. Similarly, increasing vaccine coverage to 70% was deemed not cost-effective (ICER=$22,623 (22,602 to 22,645) per DALY averted) compared to 50% coverage at a cost-effectiveness threshold of $919.11.

Under the rapid vaccination scenario, a 30% vaccine coverage strategy was even more cost-effective ICER=$-1,607 (-1,609 to -1,604) per DALY averted compared to the same coverage level under the non-rapid scenario, and hence is more cost-effective. The ICERs of 50% and 70% coverage strategies under the rapid scenario are $18,257 (18,226 to 18,287) and $44,250 (44,126 to 44,374) per DALY averted compared to 30% and 50% coverage strategies respectively and hence are not cost-effective.

### Sensitivity Analysis

Table 3 presents the univariate sensitivity analysis of under-reporting of hospitalizations and deaths, from a societal perspective. Assuming no under-reporting or adjusting the under-reporting factor to 2, results in all the scenarios having ICERs above the cost-effectiveness threshold, except the 30% coverage with a rapid deployment. On the other hand, with an under-reporting factor of 3 or 4, 30% coverage with a rapid and non-rapid vaccination scenario remained cost-saving.

Figure S6 summarizes the effects of vaccine prices and discounting rates of DALYs on the ICER. Vaccine prices, of the two parameters had the largest effect on the ICERs: leading to a 32-103% decrease and a 36-77% increase in ICERs across the different vaccination scenarios.

The one-way sensitivity analysis focusing solely on a health system’s perspective is presented in Table S3. When considering this perspective, the total costs across the vaccination strategies increase as coverage increases, as reported from a societal perspective. However, the no vaccination scenario affords the least costs ($313 million). The reported ICERs increase with increased coverage and the 30% coverage with a non-rapid and rapid vaccination pace are below the threshold: ICER=$555 (553 to 557) and $291 (290 to 295) per DALY averted respectively and considered cost-effective from a health system’s perspective.

Figure 2 represent the findings of the probabilistic sensitivity analysis from a societal perspective. The region below the cost-effectiveness threshold line and within the grey region, shows all the points that are cost-effective at a cost effectiveness threshold of $919.11. For instance, the dominance of the 30% coverage scenarios (i.e. more effectiveness at a lower cost) compared to no vaccination, was shown in 100% of the replications (i.e. 100% of the cost-effect pairs were in the southeast quadrant). Further, 100% of the replications for 50% coverage and 70% coverage strategies (both rapid and non-rapid rollout) were in the northeast quadrant (implying that these strategies were more costly but also more effective compared to the 30% and 50% coverage strategies respectively).

**Figure 2:**
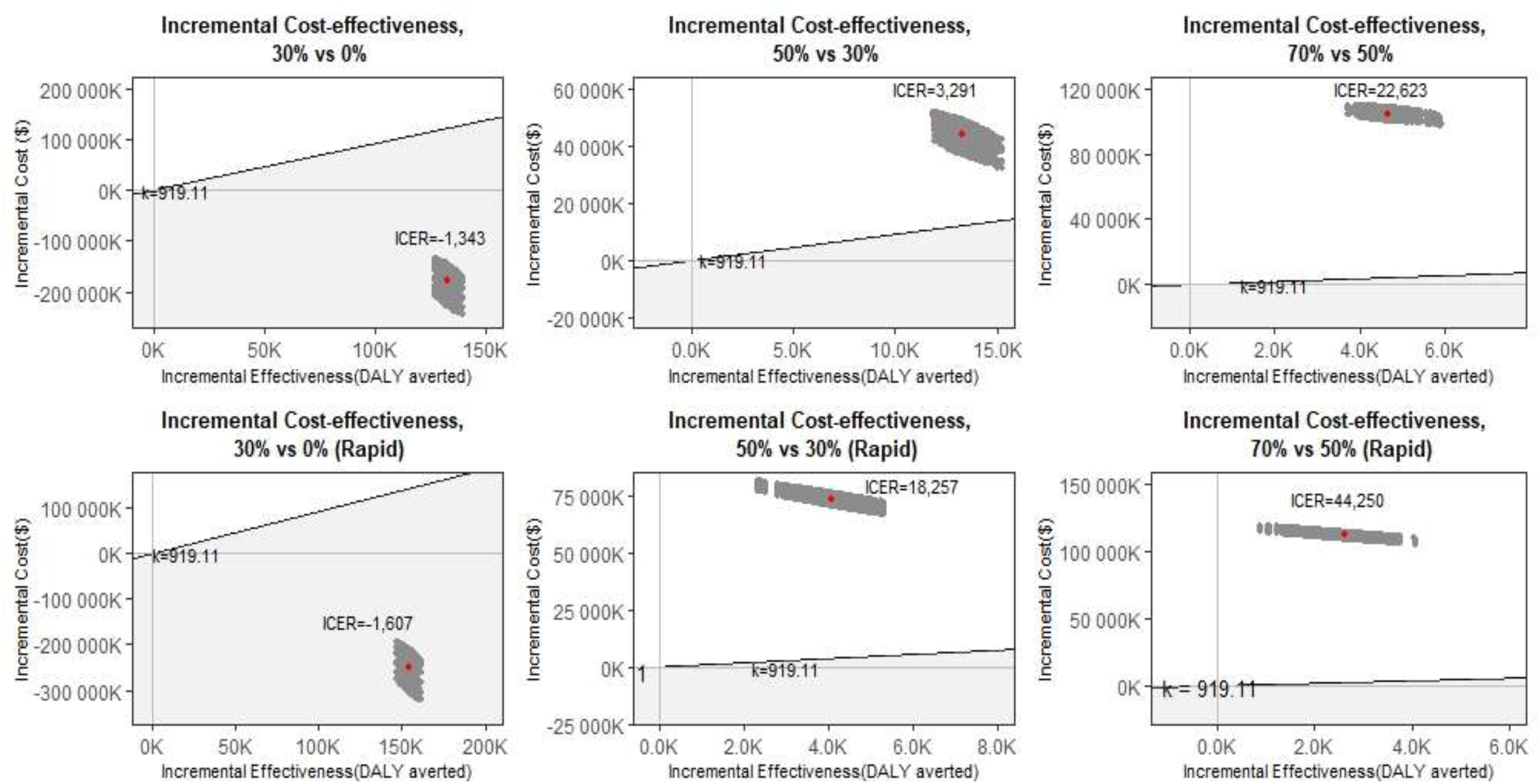
Probabilistic sensitivity analysis of different vaccination strategies from a societal perspective. The first row shows the vaccine scenarios comparisons under a non-rapid rollout pace while the second row shows the rapid roll out results. Each grey dot represents a pair of values of incremental cost and incremental effectiveness and the red point is the mean ICER points for each vaccine comparison. The grey shaded area below the diagonal cost-effectiveness threshold line (k=919.11 USD) shows the cost-effective region

Figure 3 presents the cost-effectiveness acceptability curves (CEAC) of the analysis from a societal perspective based on a range of cost-effectiveness thresholds. Under the non-rapid vaccination rollout and given a $3,300 willingness to pay threshold, the probability of the 50% coverage strategy being cost-effective compared to 30% coverage would be 0.5. Further, there was 0.5 probability that the 70% coverage in comparison to 50% coverage would be cost-effective at a threshold of $22,600 in the non-rapid deployment.

**Figure 3:**
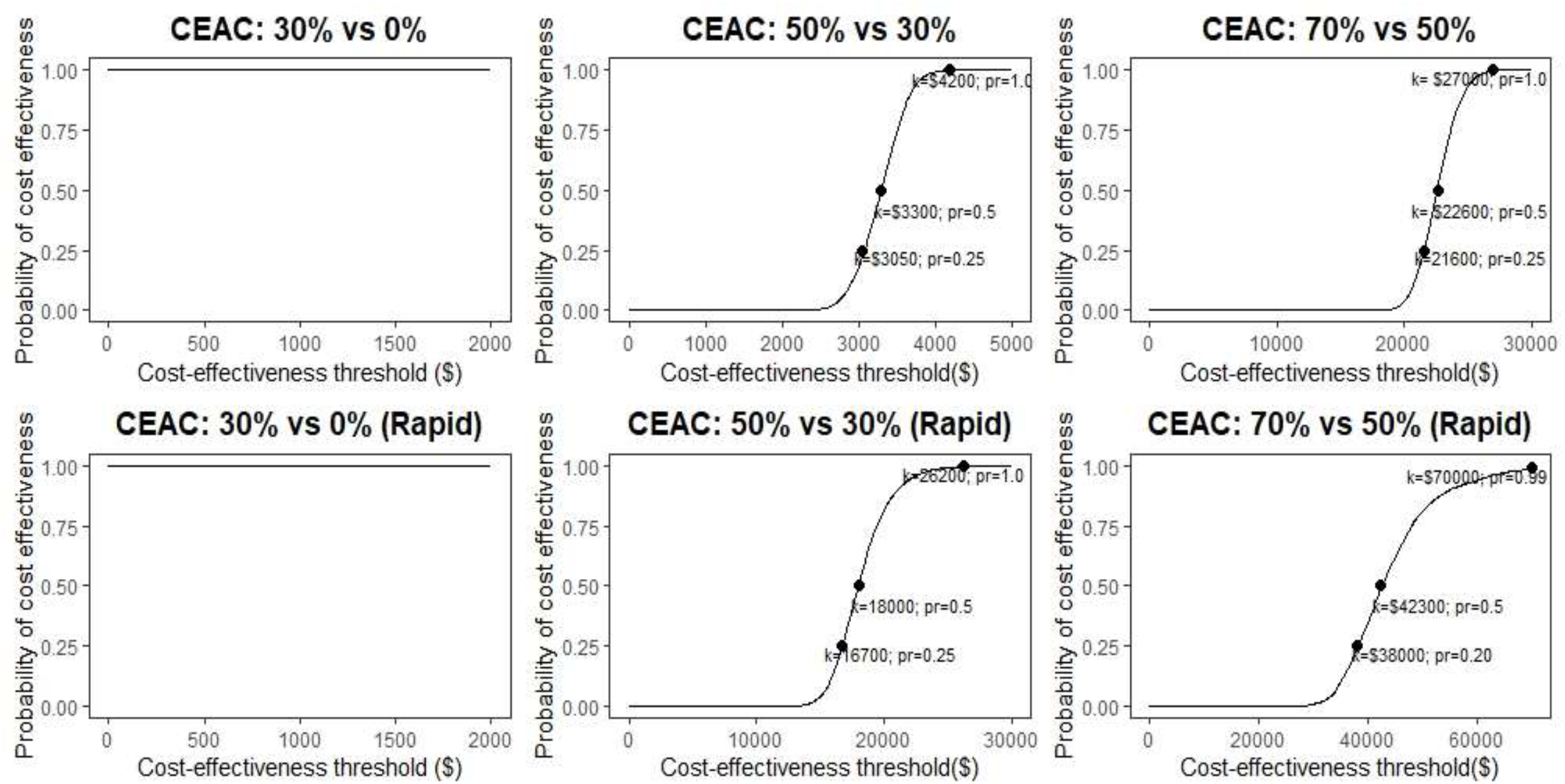
Cost-effectiveness acceptability curves showing the probability that each index scenario is cost-effective compared to the comparator over a range of cost-effectiveness thresholds (k=cost-effectiveness thresholds, pr=probability of cost-effectiveness)

## DISCUSSION

We assessed the epidemiological impact and cost-effectiveness of a range of COVID-19 vaccine deployment strategies and scenarios in Kenya. Our findings show that if Kenya had started with a full-scale vaccination programme in September 2021 and with an omicron-like variant introduced in November 2021, the deployment of COVID-19 vaccines in the Kenya population would likely avert a substantial number of cases, hospitalizations, and deaths from COVID-19. We find that a strategy to vaccinate mostly older adults (80% of those over 50y) who are at high risk of severe disease but which achieves low (30%) overall population coverage, yields the greatest reductions in severe infections and deaths per fully vaccinated adult. The marginal health benefits decrease with higher vaccine coverage levels (50% and 70%) as an increasing proportion of low risk younger adults, most with some immunity from previous infection, are vaccinated. These diminishing returns of increased coverage result in only the programme for older adults (i.e. the 30% coverage scenario) being cost effective while the expansion to younger age groups (i.e., 50% and 70%) was found not cost effective. Further, where an upsurge of SARS-CoV-2 occurs shortly after scale-up of vaccination (as modelled in this study) then deployment strategies that achieve rapid coverage of the target groups are more effective compared to slow vaccine deployment strategies.

Our findings are similar to evidence from South Africa, Madagascar, Pakistan, United Kingdom (UK), and United States of America (USA) that found vaccinating their population would decrease COVID-19 infections and deaths compared to a no vaccination scenario[8,37–40] and increasing vaccination coverage would increase the clinical benefits.[37,39] The South African study also found that a rapid vaccination roll-out pace resulted in “better” clinical outcomes (infections and deaths averted) and economic effectiveness compared to a non-rapid roll out pace.[37] The studies done in Madagascar, UK, and USA reported a greater impact when distribution of vaccines was prioritised according to the number of people of an older age in the region or among the elderly, reflecting similar findings to our study.[8,38,40] However, in contrast to the South African and Pakistan studies[37,39] who found that higher coverage scenarios had higher marginal impacts, we found that a minimal vaccine coverage of 30% of the adult Kenyan population targeting older age groups had the highest marginal impact. These differences could be explained by differences in the demographic profiles of the different populations of study. Higher population coverage with the COVID-19 vaccines have greater health impacts in countries that have higher proportions of the elderly and/or low previous exposure to COVID-19.

Using a societal perspective (that incorporates health system costs and productivity losses), we find that COVID-19 vaccination in Kenya is most cost-effective when targeted at older age groups in the population. This is because all our scenarios have the elderly covered first, and the incremental impact of increasing vaccination coverage among younger populations was less value for money. Given that the proportion of the elderly population in Kenya is low (11% of total population are aged 50 years and above),[13] targeting the COVID-19 vaccine to this vulnerable population achieves high cost-effectiveness at relatively low population-level vaccine coverage; 30% coverage of the population ensures that the maximal 80% of the older age group is vaccinated and a very low coverage of the younger age group (19%). Accounting for productivity losses improves the cost-effectiveness profile of COVID-19 vaccines, compared to when only direct health system costs are considered. For instance, for the 30% coverage scenarios with both a non-rapid and rapid deployment pace, the ICERs decreased on average by 342% and 652%, when the societal perspective was considered as opposed to the health system perspective, and as a result improving the cost-effectiveness profile. This underlines the limitations of using a narrow health system perspective that ignores broader societal costs of health system interventions. This is even more so for a vaccine deployed in a pandemic that has substantial socio-economic impacts, in addition to health impacts. These findings mirror cost-effectiveness studies of COVID-19 vaccination done in Turkey and Pakistan that found that although COVID-19 vaccination strategies were cost-effective from a health system’s perspective, they were cost-saving from a societal perspective.[39,41] This is in line with arguments from studies that estimate the public health value and impact of vaccination, which argue the need to broaden the perspectives for cost-effectiveness analysis of vaccines, as their impact is far-reaching, especially in the context of a pandemic.[42–44]

These findings have implications for COVID-19 vaccination policy in Kenya and other low-and middle-income countries (LMIC) settings with comparable demographic and COVID-19 epidemiological profiles. First, not unexpectedly, where an outbreak is imminent efforts to rapidly deploy the vaccine not only avert more cases, hospitalization, and deaths, but are also more cost-effective. By extension, had Kenya been able to deploy vaccines more rapidly, benefits would have been greater. Second, COVID-19 vaccination is likely to offer the best value for money when targeted to older age groups and possibly other vulnerable groups (such as those with risk increasing comorbidities) with high risk of severe disease and death, rather than to the whole population, in settings with overall low risk of severe disease and deaths, and high natural immunity due to previous exposure. This has several further implications. Kenya and other similar settings will achieve better health impacts and value for money with relatively small numbers of vaccines targeting the high-risk sections of the population. Against a context of constrained fiscal space for health, it is likely more prudent for Kenya and other African countries to target the vulnerable rather than whole populations. This consideration is likely to be even more relevant as African countries consider two shifts; the eligibility of children (below the age of 18 years) to COVID-19 vaccination and the transition to endemicity. If an endemic scenario will require annual vaccinations, Kenya and other African countries are unlikely to afford yearly vaccinations of their entire population. It is also apparent that such a strategy (vaccinating the entire population) is unlikely to be cost-effective, necessitating the need for Kenya and other African countries with comparable demographic and epidemic profiles to be both pragmatic and evidence-based in setting COVID-19 vaccine coverage policies and targets that are both feasible, effective, and cost-effective in their contexts (rather than replicating high income country strategies.

These results should be interpreted within the context of several limitations. First, our results are dependent on model assumptions and input parameters, as is the case with all modeling studies. We selected transmission model parameters based on published literature and available observation data. However, some data was limited, lacking, or uncertain and therefore we assumed our “best” estimate for Kenya. For example, we used estimates of vaccine effectiveness based on UK data and assume a duration of 14 days between vaccination and peak efficacy within our model structure. We noted from literature,[31–34] vaccine effectiveness varies with age, duration between vaccination and testing of efficacy, variant of infection, and the type of vaccine amongst other factors. The model does not consider the different professions of the population such as essential workers (health care workers, teachers, among others) as it focusses on age as the key risk group. However, frontline workers may be important to target since preventing infection among them lessens the potential impact on health and learning capacity. The latter might become more influential in the future with new vaccines if they are more effective in preventing re-infection and mild symptoms than current generation of vaccines. Second, sub-Saharan African countries like Kenya have notably reported lower cases and deaths compared to other countries across the globe, this could be attributed to their lower testing capacity. Hence, we assumed an under-reporting factor of 1:5 in hospitalised cases and deaths. Third, we instituted vaccination roll out near in time to the introduction of a new variant which enhances the benefit of rapid over slow roll-out. Distance between vaccine introduction and the emergence of an immune escape variant is likely to favour slower vaccine roll out. Fourth, assumptions about wanning immunity (natural and vaccine) and varying protection depending on variants affect the results. Fifth, in relation to the economic evaluation, although the cost-effectiveness analysis was conducted from a societal perspective, some costs have not been fully captured due to unavailability of data. These costs include household indirect costs incurred due to COVID-19 illness (e.g transport costs), costs as a result of long-COVID, and reduced productivity for those in the formal sector with asymptomatic/mild disease. In the latter, although we assume that they can resume work from home/places of quarantine they may have reduced productivity which is not captured in this analysis. These costs not captured in the analysis are however expected to be minimal. Fifth, the analysis assumed similar vaccine delivery costs for both rapid and non-rapid vaccination across similar coverage levels. However, it is likely that the rapid vaccination scenario may need more resources, especially cold chain equipment to hold larger batches of vaccines at a time. Sixth, the reported uncertainty of the ICER likely does not capture the full extent of the uncertainty, given the uncertainty of the costs of a yet to be established adult vaccination programme in Kenya. Lastly, the economic evaluation considers a 1.5-year time frame, potentially excluding costs and benefits of COVID-19 that may accrue over a longer period of time.

## CONCLUSION

This study contributes to the growing body of literature on the health impact and cost-effectiveness of COVID-19 vaccines. Kenya will achieve both greater marginal health impacts and better value for money if it prioritizes a targeted vaccination strategy among those at increased risk of severe disease and at a rapid rollout speed. The cost-effectiveness of the COVID-19 vaccine should be considered alongside other priority setting considerations in the Kenyan context.

## Data Availability

All code and data for the transmission model and economic evaluation analysis underlying this study is accessible at the Github repository: https://github.com/SamuelBrand1/KenyaCoVaccines.

## STATEMENTS

### Author Contribution

EB, DJN, and PB conceptualised the study. SO, JO, CO, AK, RA, MO, AA, GMW, SU, EO, LIO, CAN, KK, PA, MM, RA, WN, IMOA, JAGS, PB, and EB acquired the data. SPCB, JO, SO and CO analysed the data. SO, JO, SPCB, CO, PB, DJN, and EB were involved in the interpretation of the data and work. SO and JO wrote the first draft of the manuscript which was subsequently revised for important intellectual content by all authors. All authors read and approved the final manuscript.

## Acknowledgement

We would like to acknowledge the Health Intervention and Technology Assessment Program (HITAP) and the Africa CDC Health Economic Program on their advice on the analysis.

## Ethics approval

The KEMRI Scientific and Ethics Review Unit approved this study under KEMRI/SERU/CGMR-C/4244.

## Patient consent

Not required.

## SUPPLEMENTARY INFORMATION

### Transmission model

#### Transmission model overview

The dynamics of SARS-CoV-2 transmission in each of the 47 Kenyan counties were assumed to follow a dynamic model adapted from a previous model;[4] here we extend the (modified SEIRS type) transmission model structure to include age stratification and vaccination. The epidemiological dynamics in each county are described by the following system of differential equations:

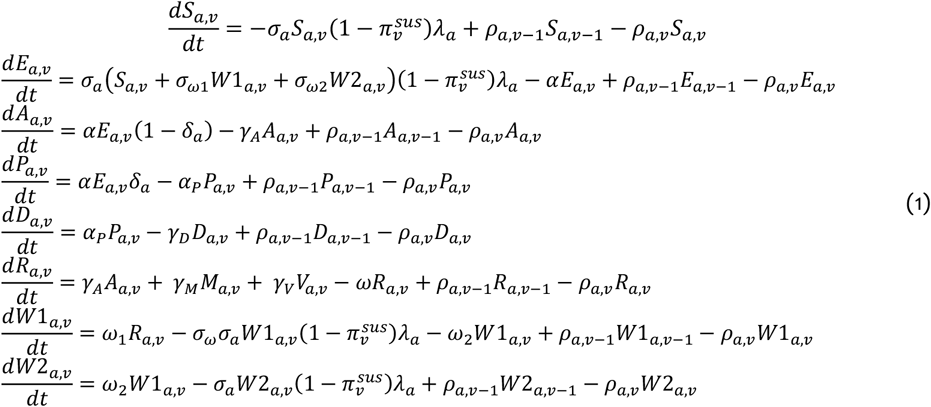

The state variables are: Susceptible (**S**), latently infected (**E**), asymptomatically infectious (**A**), pre-symptomatic infectious (**P**), symptomatic/diseased infectious (**D**), recovered and temporarily immune (**R**), and previously recovered/immune whose immunity to reinfection has waned (**W1**) before disappearing (**W2**). The indexing variables are age (*a*) and vaccination dose status (*v*), see below for further details on index variable structure. The age-specific force of infection was denoted *λ*_*a*_ (see below for details). The rate of progressing through latent uninfected stage (*α*), pre-symptomatic infectious state (*α*_*P*_), rate of loss of complete immunity in two stages (*ω*_1_, *ω*_*2*_), and the decreased susceptibility due to prior infection in stage **W1** (*σ*_*ω*_), were assumed to be identical for all age groups, vaccination statuses, prior infection events, and eventual severity of the infection episode. The recovery rates (*γ*_*A*_, *γ*_*D*_) depended on the severity of the episode, but not other factors. The baseline susceptibility per infectious contact (*σ*_*a*_) and probability of developing symptoms (*δ*_*a*_), were assumed to depend on the age of the individuals. 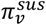 gives the effectiveness of vaccine status *v* at blocking transmission per infectious contact (see below). The per-capita rate at which a person in age group *a* and with vaccine status *v* transitioned to their next vaccine status was denoted *ρ*_*a,v*_ (see below). Figure S1 gives a visual overview of the model dynamics.

### Age structure for transmission model

In this paper we used a coarse-grained set of age indices, partly to reduce number of model compartments and thereby increase the efficiency of parameter estimation (see below), and also partly because the data used in parameter estimation did not support a finer grained age index. The six age groups corresponding to the age indices *a* = 1, …, 6 were 0-19, 20-49, 50-59, 60-69, 70-79 and 80+ year olds. When using more fine-grained age-specific data we used population weighted average values over the finer age group categories within each of the coarser six age groups used in this model to generate data. The data we used in the transmission model which was translated from a larger number of age groups to the six used in this model was as following:

- Age-and setting-specific contact rates for Kenya from Prem and Jit.[45] These are provided in 16 5-year age groups and 75+ year olds. (Fig. S2)
- Age-specific relative susceptibility to infection (*σ*_*a*_). These were accessed from UK focused modelling and analysis.[8] (Fig. S3)
- Age-specific chance of a symptomatic episode given infection (*δ*_*a*_). These were accessed from UK focused modelling and analysis.[8] (Fig. S3)

### Vaccine status and vaccination effects on transmission model dynamics and waning vaccination dynamics

We used five vaccination statuses to index individuals: unvaccinated (*v* = 1), vaccinated with one (*v* = *2*) or two doses (*v* = 3) with sufficient time elapsed since dose inoculation that maximum vaccine efficacy had been achieve (assumed to be 14 days), and two stages of waned vaccination (*v* = 4,5). The vaccination waning dynamics follow those described by Keeling et al.[16]

Vaccine status acted on the dynamics of the model in four distinct ways:

1) Decreasing the chance of infection per infectious contact by a factor 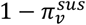 where 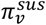 was the effectiveness of vaccine dose status *v* at reducing infection.

2) Decreasing the probability of severe disease after infection by a factor 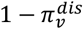 where 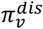 was the effectiveness of vaccine dose status *v* against severe disease.

3) Decreasing the probability of death after infection by a factor 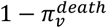 where 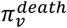 was the effectiveness of vaccine dose status *v* against death.

4) Decreasing the infectiousness of dosed infecteds by a factor 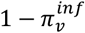 where 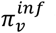 was the effectiveness of vaccine dose status *v* against transmitting infection.

We follow the “VE -> 0%” scenario from Keeling et al,[16] where the effectiveness of the vaccine against acquisition of COVID-19 and infectiousness during a COVID-19 episode eventually decreases to zero 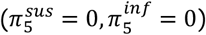, whilst the eventual effectiveness against severe disease and death decreases to 70% 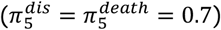. The pre-waned vaccination status (*v* = 4) has the same vaccine effectiveness as full second dose vaccination status (*v* = 3) but is used to give non-exponential waning rates over an average of 430 days for both stages of waning vaccine effectiveness. Therefore, the per-capita transition rates *ρ*_*a,v*_ in equation (1) divide into:

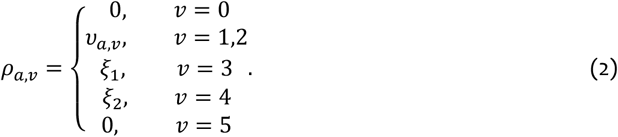

Where *υ*_*a*,1_ and *υ*_*a,2*_ are the per-capita daily rates at which individuals in age group *a* receive their first and second doses of vaccine, and *ξ*_1_ = *ξ*_*2*_ = *2*/430 per day are the waning immunity rates of vaccine effectiveness.

Although, this model choice closely follows Keeling et al,[16] it should be noted that unlike Keeling et al we assume that protection from vaccines *and* natural infection combine favorably whereas in Keeling et al it was assumed that protection from natural infection dominated vaccine protection.

### Force of infection for transmission model

We define the age-dependent force of infection (*λ*_*a*_) in three steps. First, we define the effective number of infected in each age group *b*,

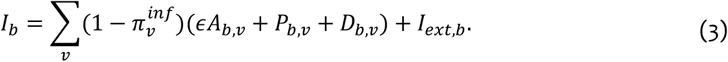

The effective number of infecteds is the total number rescaled by decreased levels of infectiousness, such as the relatively lower infectiousness of asymptomatic infecteds compared to pre- and post-symptomatic infecteds (*ϵ*). *I*_*ext,b*_ represented an external coupling with infectious people external to internal transmission dynamics. We chose *I*_*ext,b*_ such that ∑_*b*_ *I*_*ext,b*_ = 100. Second, the rate of infectious contacts from an effective infected in age group *b* to anyone in age group *a* was defined as,

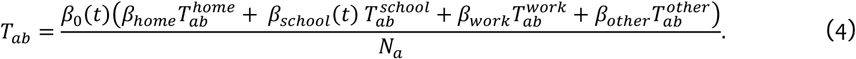

Contacts occur in any of four main settings: at home, at school, at work or in some other social setting, each of which has an age-specific contact rate matrix for Kenya 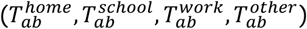 estimated by Prem and Jit.[45] Google mobility data[46] and previous epidemic modelling in Kenya [4] suggest that contact rates in Kenya had returned to approximately pre-pandemic baseline by January 2021, therefore, we treated the setting specific transmission rates per contact for at home, at work and other social setting (*β*_*home*_, *β*_*work*_, *β*_*other*_) as constant. Transmission rate per contact at schools (*β*_*school*_(*t*)) was constant during term time, but dropped to zero during Kenyan school holidays (19^th^ March – 10^th^ May, 16^th^ July – 26^th^ July, 1^st^ October – 11^th^ October, 23^rd^ December – 4^th^ January). The baseline transmission rate per contact (*β*_0_(*t*)) varied according to the SARS-CoV-2 variant frequency in the county (see below). Third, the force of infection was defined as,

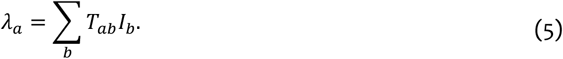

### Alpha, Beta and Delta variant frequency effect on transmission model dynamics

The Alpha, Beta and Delta variants of SARS-CoV-2 have circulated in Kenya,[47] displaying a pattern of sequential dominance. From December 2020, Alpha and Beta variants increased in frequency ([48] Fig. S2), with a complex spatial pattern of relative frequency of Alpha vs Beta within Kenya.[48] Then, from May 2021, the frequency of Delta variant increased very rapidly to complete domination across Kenya (Fig. S2).

We modelled the effect of variant frequency on the baseline transmission rate per infectious contact *β*_0_(*t*) as a sequence of strain dominations each occurring over a timescale set by a logistic growth curve,

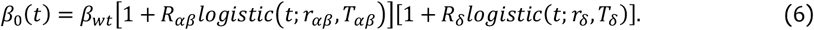

Where *logistic* 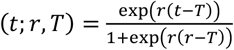. In this model, *β*_*wt*_ is the baseline transmission rate per infectious contact of the origin strain(s), called wild-type strains, circulating in Kenya before Alpha and Beta variants, (1 + *R*_*αβ*_) is the proportional change in the reproductive number for SARS-CoV-2 after domination by Alpha or Beta variant relative to wild-type strains, and (1 + *R*_*δ*_) is the proportional change in the reproductive number for SARS-CoV-2 after domination by Delta variant relative to Alpha or Beta variant. *r*_*aβ*_, *r*_*δ*_, *T*_*αβ*_, *T*_*δ*_ set the exponential rates and timing of the logistic growth curves. The implied relative frequencies of wild-type, Alpha/Beta and Delta variants over time are,

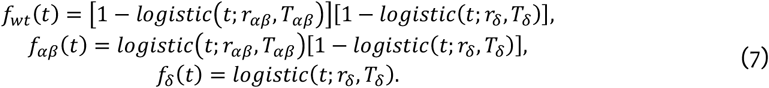

### Transmission model observables: Proportions PCR test and serology test positive

The underlying transmission of SARS-CoV-2 is not observed, rather we have access to swab tests and serological tests (positive and negative) aggregated by date, age, and county. The chance infected individuals test positive for either type of test depends on the number of days post-infection. Therefore, we coupled the dynamics cumulative infections to transmission model,

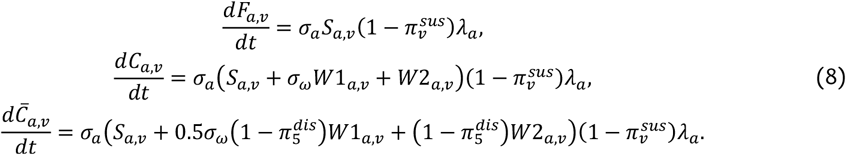

Where *F*_*a,v*_(*t*) and *C*_*a,v*_(*t*) were the cumulative number of people in the county in age group *a* and vaccine status *v* infected by time *t*, respectively split by it being their first infection episode or any infection episode. 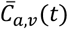 was the reinfection disease risk-weighted cumulative infection rate; that is the total infections discounted by the decreased risk of severe disease among reinfections (see below for use of this observable). In the absence of other evidence, we follow Keeling et al[16] in assuming that protection against disease due to prior infection is similar to that of vaccination. For the fully waned immunity post-natural infection state (**W2**) we assume that the protection from disease is equivalent to fully waned vaccine protection, and, for the partially waned immunity post-natural infection state (**W1**) that this protection is halved relative to **W2**.

The daily new infections, on each day *n* starting at *t* = *n* are then,

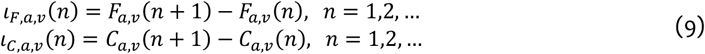

The probability that an infected individual would be determined as having been infected *τ* days after infection *if* tested by either a PCR swab test or a serology test were denoted, respectively, *Q*_*PCR*_(*τ*) and *Q*_*sero*_(*τ*). We used the same *Q*_*PCR*_ and *Q*_*sero*_ probabilities as in Brand et al 2021.[4]

By combining the underlying infection processes and the delay between infection and observability in our available data sets we find that the number of people who would test positive on each day *n* in each county with either a PCR test 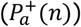, or a serology test 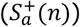, was,

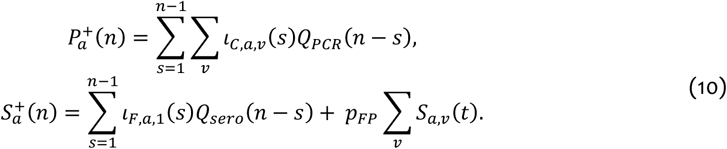

where *t* was the midpoint of day *n*.

*p*_*FP*_ was the false positive rate for the serology assay (see table S1). Underlying assumptions for equation (10) are: 1) that the PCR test is 100% specific to SARS-CoV-2, 2) that *only* the first infection contributes to the serological status of individuals, but that reinfections contribute to PCR status equally to first infections, and 3) that during the period of transmission parameter inference there were effectively zero vaccinated individuals, and therefore, all seropositivity was evidence of prior nature infection. We do not use any serological data from Kenya after May 2021 when the vaccination rate was very low in Kenya, and, therefore, vaccinations have negligible effect on prevalence of SARS-CoV-2 specific antibodies.

The number of people PCR positive is not observed directly, but rather test positive and negative swab test samples. We consider the *proportion* of these daily samples that are positive to be a potentially biased sample of the true underlying proportion that would be PCR-positive if everyone was tested 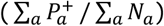. Therefore, we model the expected proportion PCR test positive (over all age groups) on day *n* as,

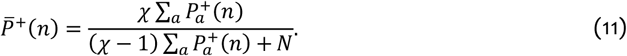

Where *𝒳* is an observed swab sample bias parameter, where *𝒳* = 1 indicates unbiased sampling, *𝒳* < 1 indicates bias in favour finding PCR negative individuals (i.e., 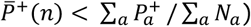), and *𝒳* > 1 indicates bias in favour of finding PCR positive individuals (i.e., 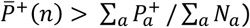).

### Clinical outcome model

#### Clinical outcomes of SARS-CoV-2 infections

Severe infections eventually lead to a clinical outcome. We consider three possibilities in this model:

1. Deadly outcome. Deadly infected individuals die after a delay period defined by the probability distribution *fμ*. Death, conditional on infection, occurs with probability 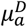.
2. Critical outcome. Critically infected individuals require a stay in ICU for a duration defined by the probability distribution *f*_*ICU*_, then move to a general ward in a hospital or health facility, where they stay for a duration defined by the probability duration *f*_*hosp*_. Critical disease, conditional on infection, occurs with probability 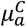.
3. Severe outcome. Severely infected individuals require a stay in a general ward in a hospital or health facility, where they stay for a duration defined by the probability duration *f*_*hosp*_. Severe disease, conditional on infection, occurs with probability 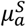.

### Clinical outcome model observables: Reported incidence of deaths, occupancy of general wards and Intensive care units

#### Incidence rate of clinical outcomes

The lag between infection and needing treatment, for those infected individuals who die, was defined as the convolution of two time-duration distributions:

1. The duration of time between infection and symptoms (days), which we assumed was distributed *LogNormal* 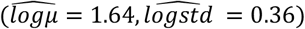.[49]

2. The duration of time between initial symptoms and severe symptoms (days), sufficient to seek hospitalisation, which we assumed was distributed *U*(1,5).[50]

We discretized the two distributions to give probability functions *f*_*IS*_ for the number of days between infection and symptoms, and *f*_*SH*_for the number of days between symptom onset and severe symptom onset. The probability function for the (discrete) number of days between infection and severe or critical disease, for those who died, *f*_*dis*_, was given as a discrete convolution over these probability mass functions:

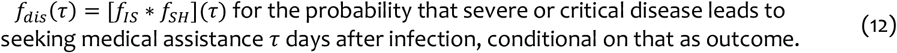

We use this delay distribution to give a rate of people in age group *a* requiring medical treatment on day *n* up to some unknown age-dependent and variant-dependent risk-factor, which will fit against available data on reported severe, critical and deadly outcomes,

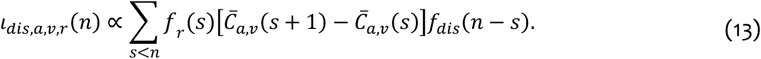

Where *f*_*r*_(*s*) is the relative frequency of variant *r* on day *s* (see equation (7)). Note that in equation (13) the reduction in risk due to reinfection is already accounted for (see equation (8)).

#### Observation of incidence of deadly outcome of infection

There is likely to be under-reporting of deaths due to COVID-19 in Kenya.[35,51] In this paper, we don’t have sufficient data to estimate the true level of under-reporting of deaths in Kenya. However, by assuming that the age-dependent risk of death after infection with SARS-CoV-2 is the same in every county, we can estimate county-specific under-reporting/change in risk *relative* to the capital Nairobi. Concretely, we model the expected number of observed deaths on each day *n*, in each age group *a*, and each county *c*, as,

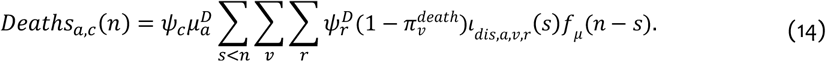

Where *ψ*_*c*_ is the county-specific under-reporting rate with 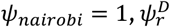 is the relative risk of death by variant with 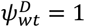, and the duration of time between needing treatment and death had probability distribution *f*_*μ*_.

#### Observation of hospital and ICU occupancy due to severe or critical infections

We don’t have access to reports on the incidence of severe and critical cases arriving at hospitals/health facilities and ICUs; the relevant observables from the clinical outcome model are occupancies of patients overall in Kenya by setting rather than arrival of patients at those settings.

The probability distributions for length of stay in either hospital/health facility or ICU are denoted as *f*_*hosp*_ and *f*_*ICU*_, implying upper distribution functions *Q*_*hosp*_(*n*) = ∑_*s*>*n*_ *f*_*hosp*_(*s*) and *Q*_*ICU*_(*n*) = ∑_*s*>*n*_ *f*_*ICU*_(*s*) for the probability that a stay in hospital or ICU is longer than *n* days. The expected total number of people in intensive care units on day *n* is,

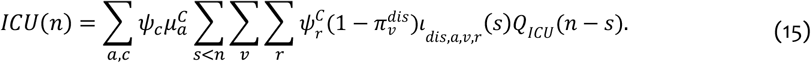

Where 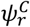 is the relative risk of critical disease by variant with 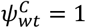. After a critical case has completed a stay in an ICU, we model them as having a stay in a general ward with the same distribution of length as per a severe case admitted to a general ward (without a stay in ICU). The upper distribution function for the whole stay in ICU *and* general ward is then *Q*_*ICUH*_(*n*) = ∑_*s*>*n*_[*f*_*ICU*_ ∗ *f*_*hosp*_](*s*), giving the expected number of critical cases in either ICU or general ward as,

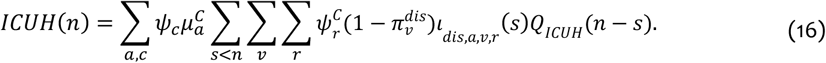

The expected number of patients occupying general wards is the addition of severe cases who have been admitted directly to general wards, and critical cases who have completed their stay in ICU and are now in general wards,

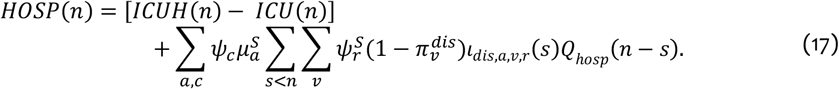

Where 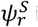 is the relative risk of severe disease by variant with 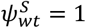.

### Parameter Inference

In this work, we make inferences on two groups of parameters:

1. The parameters of the transmission model in [equations (1-5)], and the bias parameter (*𝒳*) for the observed versus actual proportion PCR positive in daily swab test [equation (11)].

2. The parameters of the clinical outcome model [equations (12-15)]: the relative under-reporting rate by Kenyan county *ψ*_*c*_, the age-dependent clinical outcome probabilities 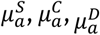, and the variant specific clinical outcome probabilities 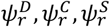.

For the transmission model parameters, we used Bayesian inference to infer a joint posterior distribution for the parameters for each county. For the clinical outcome model, we inferred parameters by minimizing the divergence between model prediction of the observables [equations (14-15) and (17)] and actual reporting, under the assumption that the rate of people arriving for medical treatment, up to the unknown risk factors [equation (13)], was that implied by the posterior mean prediction implied by the Bayesian inference of the transmission model parameters.

A challenge with using the linelist data in Kenya for inference of transmission was that the metadata concerning the reason for receiving a swab test, the levels of symptoms of people who tested positive, and their healthcare outcomes were often missing. Overall, more than 90% of the people who tested positive in Kenya, and for whom we have a description of their symptoms, reported no symptoms (asymptomatic). Therefore, unlike model-based inference for COVID-19 transmission in high-income countries we didn’t use severe outcomes such as hospitalization or death as data sources for inference, e.g.,[9,52,53] because this data was unreliable. Instead, we concentrated on fitting to the proportion positive of daily swabs test and serological tests jointly with detection rate of cases (see **Transmission model observables: Proportions PCR test and serology test positive** above). It should be noted that this meant that we didn’t use age-specific PCR test data, but rather fitted to the aggregate proportion positive over all age groups. However, we did use age-specific seroprevalence data.

We describe the three main ingredients for our Bayesian approach below: 1) the log-likelihood function for the data given a set of parameters, 2) the county-specific hierarchy of prior distributions for the parameters, and, 3) the Markov-chain Monte Carlo method used to draw parameter sets from the posterior distribution.

### Bayesian inference of transmission model parameters

#### Data and log-likelihood function for transmission model

Given the daily PCR and serology data for a county,

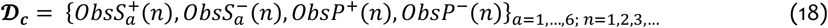

The log-likelihood function for the unknown transmission parameters (*θ*_*TM*_) in that county was,

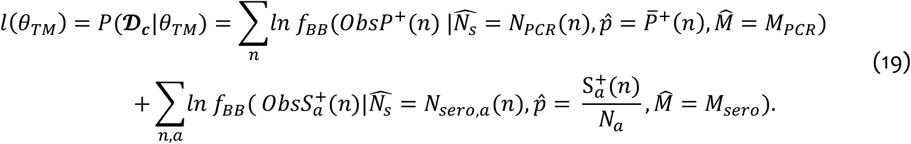

Where 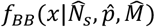 is the probability function for a Beta-binomial with sample size 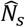, expected proportion of successes 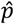, and effective sample size 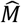. This was a convenient reparameterization of the Beta-binomial model under the transformation 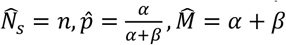 from the typical (*n, α, β*) parameterization. *N*_*PCR*_(*n*) and *N*_*sero,a*_(*n*) were the total number of samples (positive and negative) of, respectively, PCR test and serological tests on day *n*, in age group *a*. 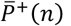 was derived from the transmission model for each day *n* as per above [equations (10-11)]. *M*_*PCR*_ and *M*_*sero*_ were fixed from a previous modelling study.[4] The first day where samples were included in the log-likelihood calculation was 1st January 2021.

#### Initial conditions and model simulation

We fit to data from 1^st^ January 2021, however, because PCR cases and serological detection are lagged indicators of infections weeks previously, we start the model simulation in each county on 1^st^ December 2020 and use the first month of simulation to allow the simulation to converge onto the epidemic dynamics. Simulation of the model was done by solving the ODE system [equation (1)] forwards from the county-specific initial conditions, with county-specific parameter configuration, using an explicit/implicit switching solver provided by the **DifferentialEquations.jl** Julia programming language package.[54]

We reduced the number of unknown parameters for the initial state of the epidemic model by considering only the overall latent infected numbers (*E*_0_) and a scale factor on the proportion exposed to COVID (i.e. in *R*/*W*1/*W2* epidemiological compartments) relative to a cross-sectional survey done in Nairobi in mid-November 2020[55] which we denote *τ*. We fix the initial removed and waned immunity numbers as

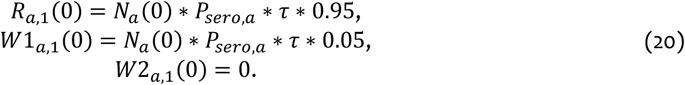

Where *P*_*sero,a*_ was the raw (non-test sensitivity adjusted) seroprevalence estimate for Nairobi in the cross-sectional survey,[55] and *τ* was an adjustment factor which was added to the set of parameters to be inferred in the set *θ*_*TM*_. Note that we are assuming that 5% of the previously exposed population have lost complete immunity to reinfection by 1^st^ December 2020, and that no previously exposed people had completely lost immunity to reinfection. The adjustment factor *τ* allowed the model flexibility to represent counties with lower seroprevalence data in 2021 as having had a smaller initially exposed fraction compared to Nairobi, whilst also allowing upwards adjustment to account for the fact that the second wave of cases in Kenya occurred during the Nairobi cross-sectional study.

The age-specific numbers of initially latent infected people were derived from the next-generation matrix *K*(*P*_*sero*,1_, …, *P*_*sero*,6_, *τ, β*_0_, *β*_*school*_, *β*_*home*_, *β*_*other*_, *β*_*work*_), where we have made explicit the parameters being inferred that the next-generation matrix depends upon and its explicit dependence on the baseline seroprevalence estimates. The eigenvector ***ν***, normalized such that |***ν***|_1_ = 1, associated with the leading eigenvalue of *K*, represented the expected distribution of new infections across age groups. Therefore, we specified

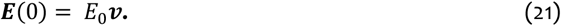

Where ***E***(0) = [*E*_1,0_(0), …, *E*_6,0_(0)]^T^. The rest of the initial variables were specified as being dependent of the flow out of the latent infected state,

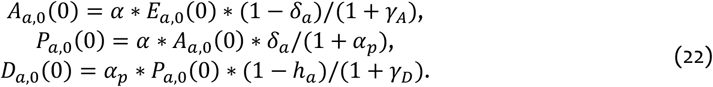

In every county, every individual in the model was initially unvaccinated.

Priors. In every county we used the following priors for parameter inference:

- 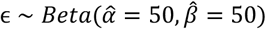
- 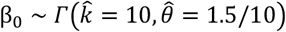
- 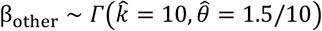
- 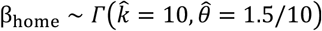
- 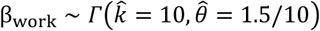
- 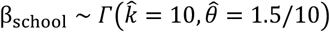
- 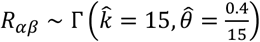
- 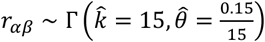
- 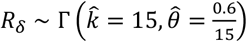
- 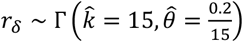
- 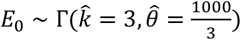
- 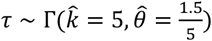

The prior for the PCR observation bias parameter *𝒳*, differed between counties (see below). For Nairobi and Mombasa we used a prior:

- 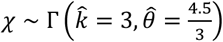

#### MCMC draws

We used Hamiltonian MCMC with NUTS[56,57] to perform Bayesian inference by drawing 2,000 samples from the posterior distribution,

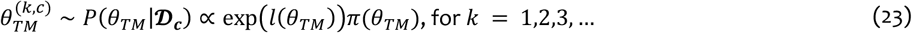

for each county using the NUTS-HMC sampler implemented by the Julia language package *dynamicHMC*.*jl*. The HMC method required a log-likelihood gradient, ∇_*θ*_(*l* + *π*), which, for our use-case of an ODE system with a comparative low number of parameters (<100 parameters), was most efficiently supplied by forward-mode automatic differentiation implemented by the package *ForwardDiff*.*jl*. The MCMC chain converged for each county (all MCMC chains and MCMC diagnostics can be accessed through the linked open code repository: https://github.com/SamuelBrand1/KenyaCoVaccines. The posterior mean (and 95% CIs) for each parameter can also be found in the open code repository.

#### Approximate county-specific hierarchical model for PCR observation bias

The serological data is important for our inference because it gives information about the proportion of each age group infected at different time points, and, therefore, allows the PCR observation bias parameter (*𝒳*) to be identifiable. However, the amount of serological data differs from county to county. To allow cross-inference between counties for the bias parameter we assumed that 1) Nairobi and Mombasa were sufficiently distinct from other counties that the inferred bias parameter for these city/counties was not relevant to other counties, and 2) the other 45 counties had a bias parameter drawn from a common distribution, *𝒳*_*c*_ ∼ Γ(*k*_*𝒳*_, *θ*_*𝒳*_), where *k*_*𝒳*_ and *θ*_*𝒳*_ are the hyperparameters of this hierarchical model. This reflected our underlying belief that despite regional variations in transmission, the observation of data would be similar in all counties outside of the main two urban hubs.

A fully Bayesian approach to inference would involve including {*𝒳*_*c*_}_*c*_ and the hyperparameters *k*_*𝒳*_, *θ*_*𝒳*_ within a joint log-likelihood over all Kenyan counties (except Nairobi and Mombasa). However, to accelerate inference we used an approximation to this hierarchical model. The 9 counties with the most amount of serological data available, apart from Nairobi and Mombasa, were **Embu, Kilifi, Kisii, Kisumu, Kwale, Nakuru, Nyeri, Siaya**, and **Uasin Gishu**. We performed MCMC draws for each of these counties using a prior 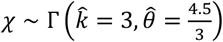, which gathered a set of MCMC draws for *𝒳* from the posterior distribution for each county *c*, {*𝒳*^(*k,c*)^}_*k*=1,…,2000_ ∼ *P*(*𝒳*|**𝒟**_***c***_). We then approximated maximum a-posteriori (MAP) estimates for the hyperparameters *k*_*𝒳*_, *θ*_*𝒳*_ using,

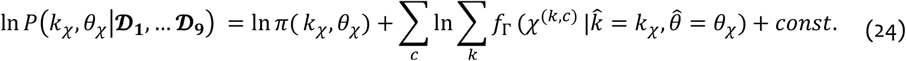

Where *f*_Γ_ is the density function of a Gamma distribution, and ln *π*(*k*_*𝒳*_, *θ*_*𝒳*_) was the log-prior for the hyper-parameters. The hyper-priors used were:

- 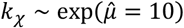
- 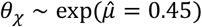

The MAP estimation is done under the distributional assumption that the observed outcome data was distributed negative binomially with the same mean as the posterior predictive mean value (that is averaged over the posterior predictive distribution for the infection process), and a negative binomial clustering factor inferred jointly with the risk factors.

We then could use the 9 non-city counties with the most serological data to create MAP estimates 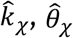 by maximizing equation (24). Equation (24) represented an approximation where we treated the

MCMC draws of the *𝒳* parameter as “data” for making inference on the hyper-parameters despite using a different prior to generate the MCMC samples.

The second approximation is that for the 36 other counties that were not Nairobi, Mombasa, or one of the 9 listed above, we used a prior for *𝒳* generated from these MAP estimates

- 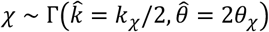

The reason for the scaling of 2 was to increase the prior variance for *𝒳* to reflect that we are approximating a hierarchical model.

### Minimum divergence estimates for the infection outcome model

After performing MCMC we were able to estimate the posterior mean for the rate of diseased incidence, up to a proportionality with unknown risk factors *E*[*l*_*dis,a,v,r*_(*n*)|**𝒟**], for each day *n*, age group *a*, variant *v*, and county *c*, by solving the ODE system [equation (1)] for each set of transmission model parameters drawn from the MCMC and using equation (13). We can then define the posterior expected number of deaths 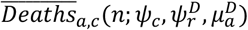 by replacing the parameter-specific diseased incidence rate in equation (14) with its posterior mean. We define the divergence due to a choice of age dependent mortality rates 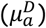, relative reporting rate (*ψ*_*c*_), relative variant-specific risk of death 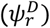 and clustering factor *a*_*D*_,

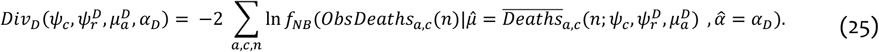

Where 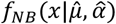 is the probability function for a negative binomial with mean 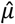 and clustering factor 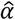, and *ObsDeaths*_*a,c*_(*n*) are the daily reported deaths in each age group, each county and on each day. We found a minimum point for equation (25), which we used as estimators, 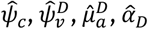 (Figs. S2, S3).

The ICU occupancy data was not available broken down by age or county, therefore, we assumed that the risk of reported critical disease was proportional to the risk of death for everyone and focused on fitting the relative risk of critical disease vs death 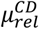. We defined the divergence between model prediction of ICU occupancy and observed occupancies due to relative risk of critical disease vs death 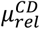 variant specific risk of critical disease 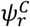 and clustering factor *a*_*c*_,

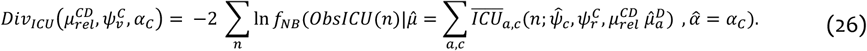

Where *ObsICU*(*n*) was the reported Kenyan National ICU occupancy on day *n*, and 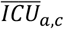 is the posterior mean value for the ICU occupancy with COVID. The minimum point for equation (26) gave estimators, 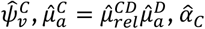 (Fig. S2).

The general ward occupancy data was also not available broken down by age or county, therefore, we assumed that the risk of reported severe disease was proportional to the risk of death for everyone and focused on fitting the relative risk of severe disease vs death 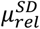. We defined the divergence between model prediction of general ward occupancy and observed occupancies due to relative risk of severe disease vs death 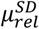, variant specific risk of severe disease 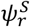, and clustering factor *a*_*S*_,

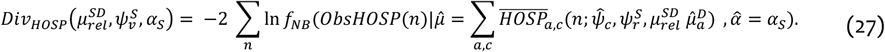

Where *ObsHOSP*(*n*) was the reported Kenyan National general ward occupancy with COVID on day *n*, and 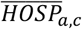 is the posterior mean value for the ICU occupancy, with the contribution from patients arriving into general wards from ICU already calculated using the minimum divergence estimates from equation (26). The minimum point for equation (27) gave estimators, 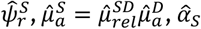 (Fig. S2).

### Vaccine scenario projections and immune-escape variant

#### Vaccination rates

We considered 7 vaccine rollout scenarios starting from 1^st^ September 2021: No vaccination (baseline), 30%, 50%, 70% target coverage of Kenyan over 18s with either an 18 month or 6-month (rapid) time scale (Table 1). In each case we assumed that:

1. The number of vaccines deployed in each county each day was constant over the rollout.

2. Second dose followed first dose after a 56 day lag.

3. Over 50 year olds were offered the vaccine first, but demand saturated at 80% coverage among over 50 year olds (age groups 3-6 in the model) and afterwards the vaccine was offered to 18-49 year olds (age group 2).

4. Vaccines were deployed pro-rata across all disease/infection states; that is there was no dependence on past infection history in seeking vaccines.

Mathematically, this corresponds to this choice for the per-capita vaccination rate (equation (2)) for the first dose, that is pro-rata distribution among all unvaccinated groups in stages by age,

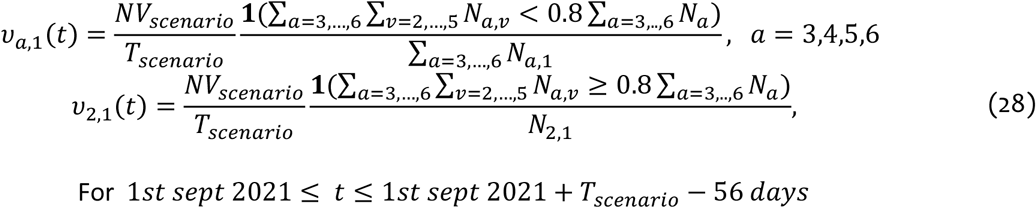

Where *N*_*a,v*_ is the county population by age and vaccine status (summed over disease/infection status). **1**(*·*) is an indicator function enforcing that the vaccination rate among over 50s (age groups 3-6) drops to zero at an 80% coverage. *NV*_*scenario*_ is the number of doses implied by the scenario target coverage, and *T*_*scenario*_ is the time in days over which this target coverage is to be achieved in the scenario. The second dose per capita rate is like the first dose but with doses distributed among people who have had their first dose,

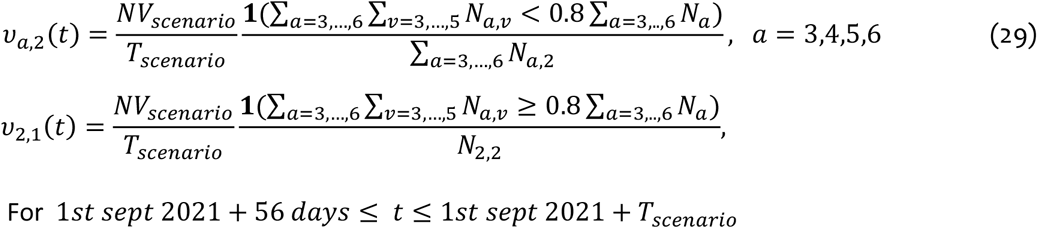

#### Uncertainty propagation in vaccination scenarios

In this study we use a mixture of full Bayesian inference, for transmission model parameters, that were specific to each Kenyan county, and minimum divergence estimators, for outcome model parameters, that were specific to Kenyan counties, e.g. the county-specific reporting/disease rate relative to Nairobi *ψ*_*c*_, or were specific to particular age groups, e.g. the baseline risk of death per infection in each age group 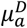.

When generating scenario projections for all Kenya, we solved the transmission model (equation (1)) for each of the 47 Kenyan counties and for each of the 2000 MCMC draws of county-specific transmission model parameters. To match the 2000 MCMC draws for transmission parameters per county we drew 2000 replicates of the vaccine effectiveness from reported ranges first and second dose AstraZeneca effectiveness against Delta variant. This generated 2000 expected daily reported death incidence, ICU occupancy and general ward occupancy for each county (equations (14-15), (17) for each county) and for each of the 7 vaccine rollout scenarios.

To account for (1) uncertainty in transmission parameters, (2) unpredictability in reporting, and (3) uncertainty in vaccine effectiveness, the prediction intervals for Kenya as a total were calculated by

1. Augmenting the 2000 projections of expected daily reported observables per day, per age group, and per county into a single group of 2000 projections of expected daily reported observables per day, and per age group by summing across counties.

2. Converting from *expected* daily reported observables to *random* instances by sampling from the negative binomial distribution that minimized divergence between actual observed data and the model projections (equations (25-27)) for each day of the 2000 projections.

3. Presenting the daily ensemble average (and 95% ensemble prediction intervals) across the 2000 randomized projections.

#### Modelling immune escape variant

In this paper we consider an immune escape variant which reduces protection against reinfection by 50% across natural immunity and vaccine protection, and spreads faster through the population with a 30% reduced generation time but is otherwise epidemiologically like the Delta variant. Concretely, we implemented this by assuming that the immune escape variant arrived in Kenya on 15^th^ November 2021, during a period of low infections for other variants, and applying a set of instantaneous effects:

- Relative immune escape variant frequency becomes 100% in all Kenyan counties, reflecting rapid dominance of invading variant.
- All transmission rate parameters (e.g. *β*_*0*_, *a, a*_*P*_, *γ*_*D*_, *γ*_*A*_) increase by a factor 1/0.7.
- 50% of people in fully or partially immune (post-natural infection) categories (**R, W1)** transition instantly to completely waned immunity to reinfection (**W2**).
- Vaccine effectiveness against reinfection and reduction of infectiousness 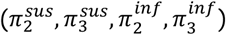 decrease by 50%.

**Figure S1:**
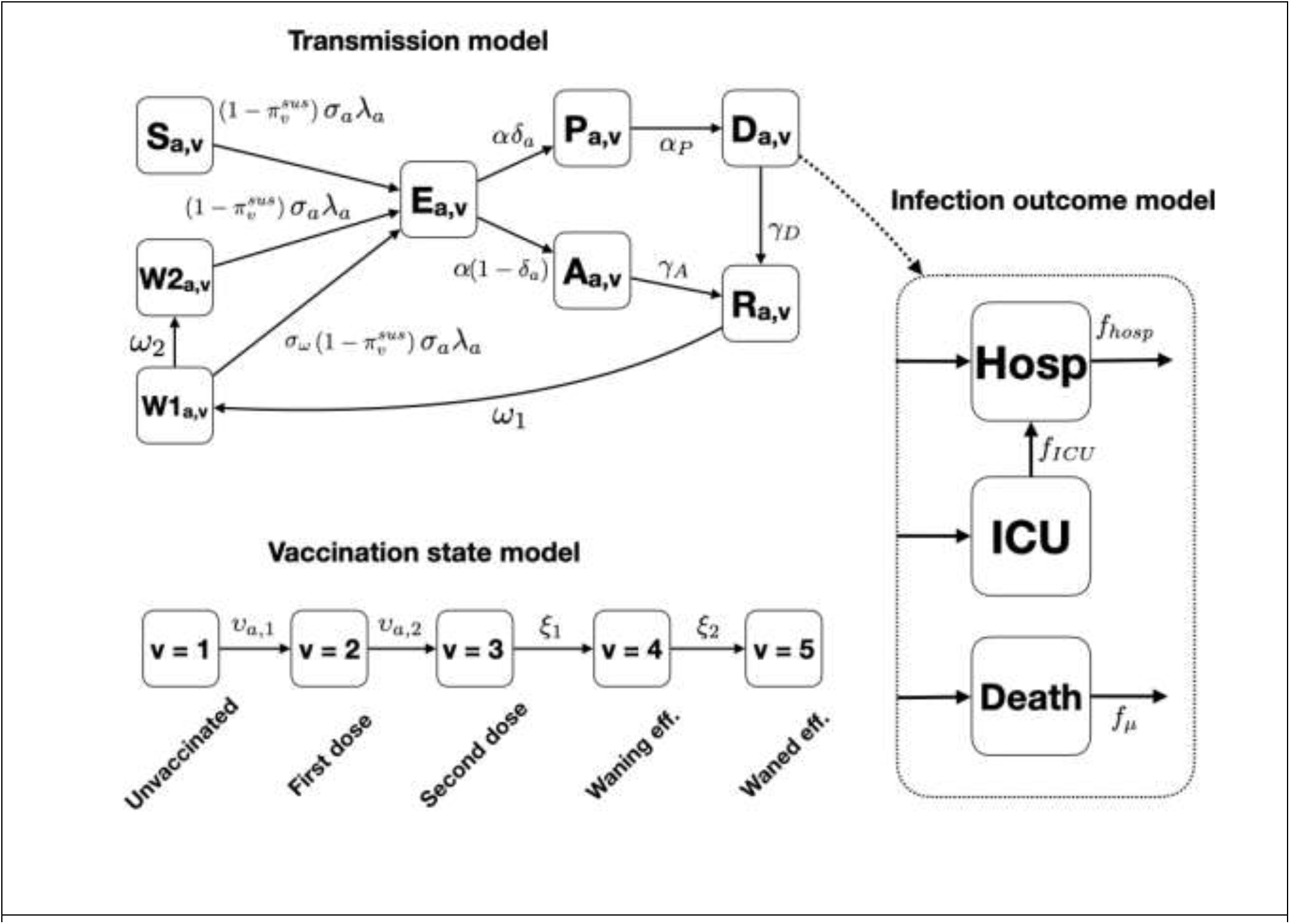
Schematic diagrams of the main modelling components. **Top left**: The transmission model for each Kenyan county follows an essentially SEIR type pattern with separate categories for symptomatic and asymptomatic infection, and two categories of waning natural immunity. **Right**: The infection outcome model for a fraction of infections that are either deadly, critical (lead to a period in ICU followed by a period in general wards), or severe (lead to a period in general wards). **Bottom left:** Vaccination state model. Transitions indicate per capita inoculation rates with first and second doses of vaccine followed by two categories of waning vaccine-induced immunity.

**Fig S2:**
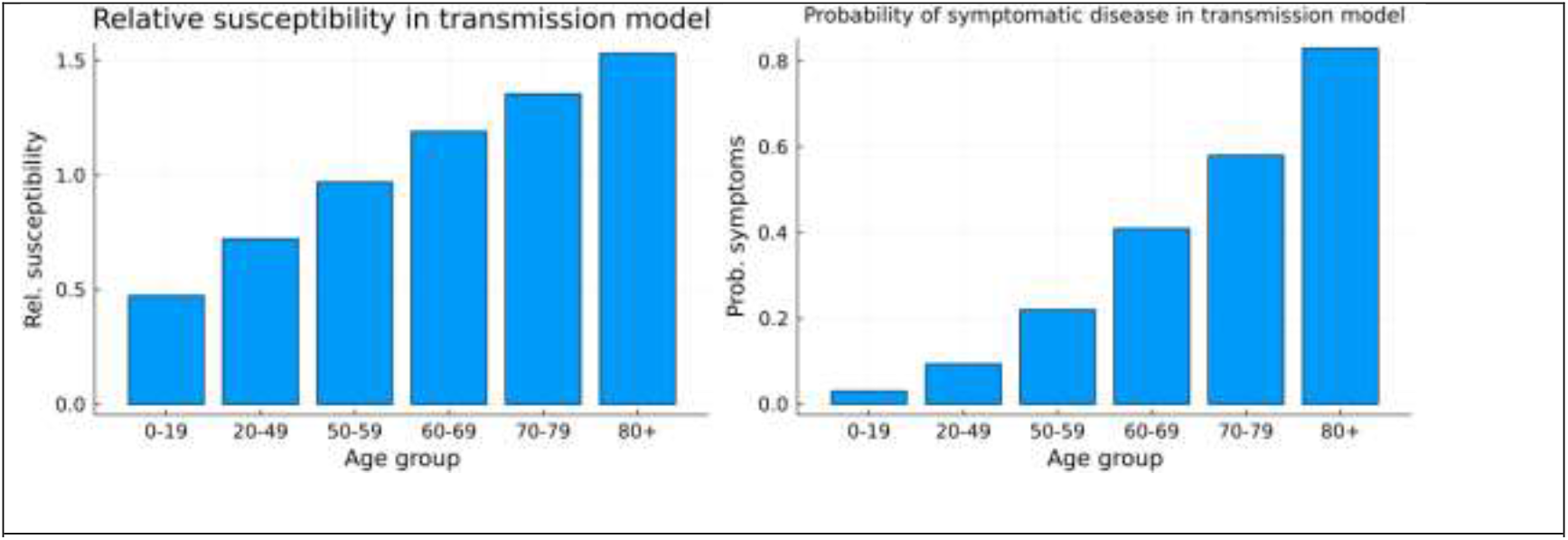
Fixed age-dependent rates. **Left:** Relative susceptibility per infectious contact by age group. **Right:** Probability of a symptomatic episode per infection for each age group.

**Fig S3.**
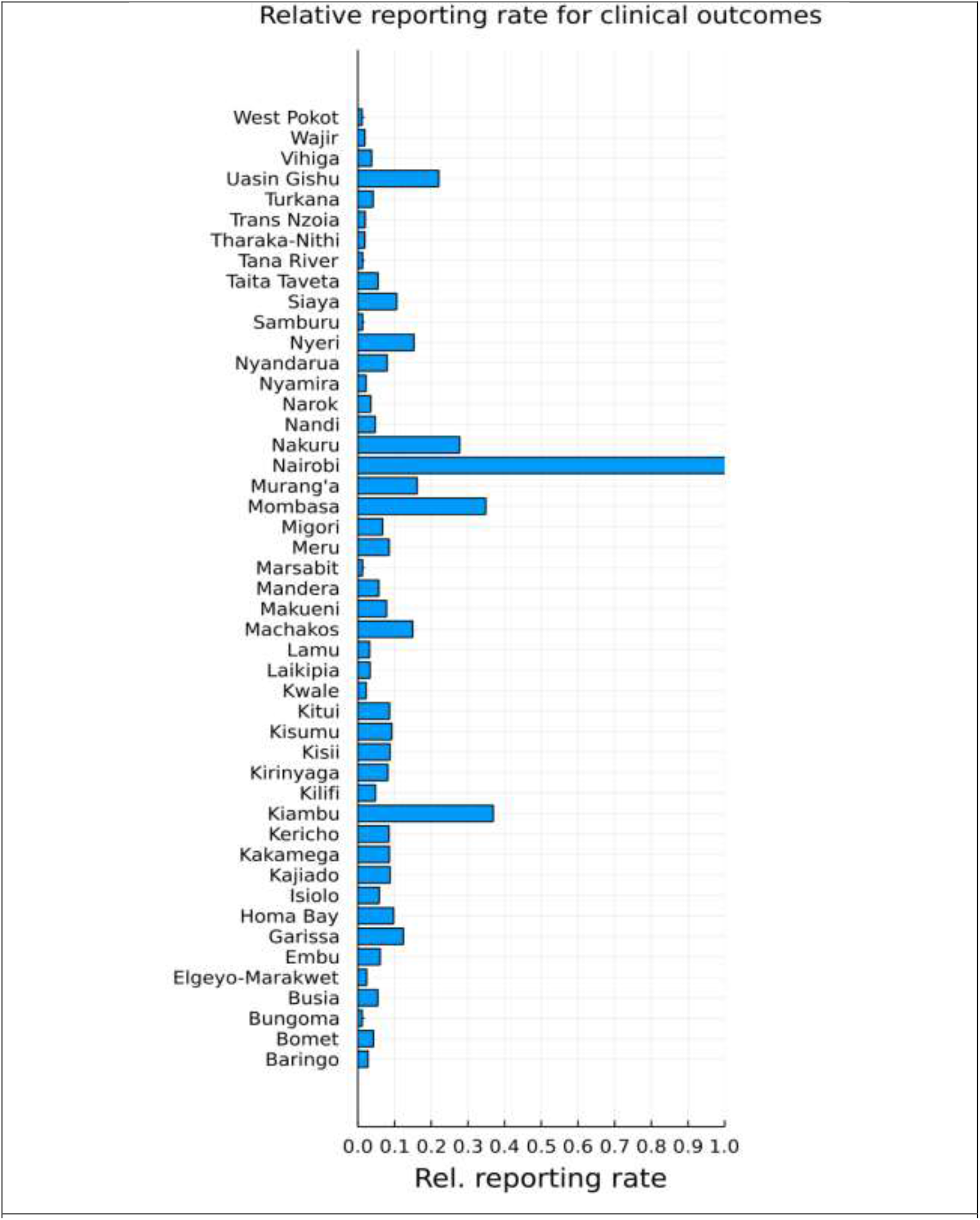
Inferred county specific reporting/disease rates relative to Nairobi (*ψ*_*c*_).

**Fig S4.**
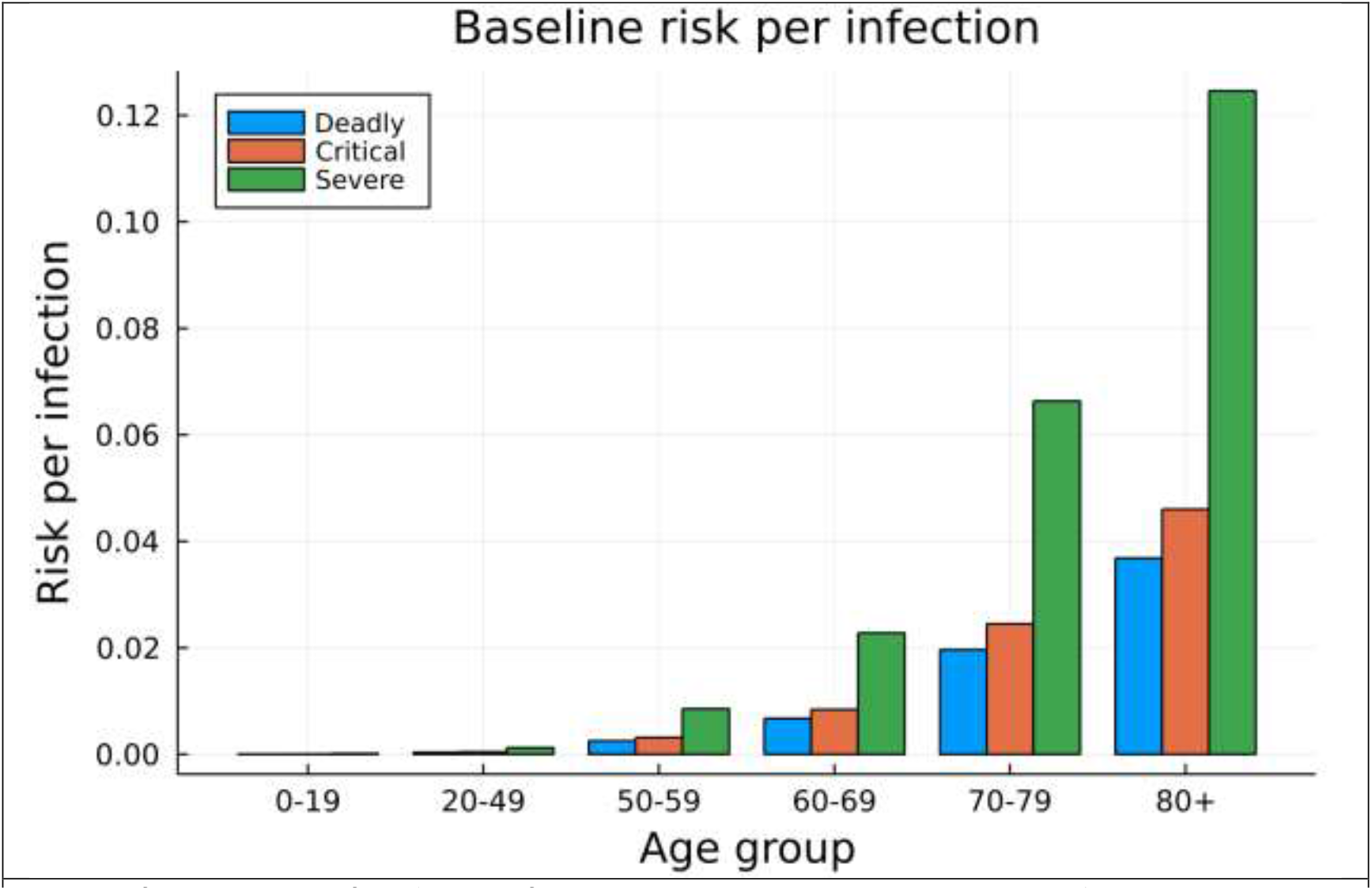
Inferred age-specific risks per infection in unvaccinated and naïve individuals.

**Table S1:** Transmission model parameters

| Transmission model parameters and variables | Value |
| --- | --- |
| **State Variables** |  |
| Number of susceptible people at time $t$, $S(t)$ | Dynamic |
| Number of latently infected people at time $t$, $E(t)$ | Dynamic |
| Number of infectious people at time $t$, $I(t)$ | Dynamic |
| Number of recovered and immune people at time $t$, $R(t)$ | Dynamic |
| Number of people who have lost natural immunity at time $t$, $W(t)$ | Dynamic |
| Cumulative number of primary infections at time $t$, $(F(t))$ | Dynamic |
| Cumulative number of all infections at time $t$, $C(t)$ | Dynamic |
| **Transmission model parameters** |  |
| Number of latently infected people at time $t=0$, 1<sup>st</sup> January 2021, by age group, $E_a(0)$, $a = 1, 2, \dots, 6$ | **inferred from data.** |
| Background transmission, $\beta_0$ | **Inferred from data** |
| Scaling from social to effective contacts at home, $\beta_{home}$, school, $\beta_{school}$, work, $\beta_{work}$ and other places, $\beta_{other}$ | **Inferred from data** |
| Maximum increase in transmission due to the introduction of the alpha and beta variants, $L_{\alpha\beta}$, or the delta variant, $L_{\delta}$ | **Inferred from data** |
| Growth rate of the alpha and beta variants, $\kappa_{\alpha\beta}$, or the delta variant, $\kappa_{\delta}$ | **Inferred from data** |
| Midpoint of the sigmoid growth curve for alpha and beta variants, $t_{0_{\alpha\beta}}$, or the delta variant, $t_{0_{\delta}}$ | **Inferred from data** |
| A factor that translates the initial (1<sup>st</sup> Jan -10<sup>th</sup> Mar 2021) seroprevalence by age group to initial attack rates, $\tau$ | **Inferred from data** |
| Infectious period, $1/\gamma$ | 2.4 days. Chosen to recreate a serial interval of 5.5days [58] |
| Latent period, $1/\sigma$ | 3.1 days. The mean incubation period [49] was reduced by two days of pre-symptomatic transmission [59] to give a latency period. |
| Mean period of complete protection after a natural infection, $1/\omega$ | 180 days, point estimate based on reinfection studies [60–63] |
| Relative susceptibility compared to naïve individuals after the loss of complete protection after the first infection, $\sigma_{\omega}$ | $\sigma_{\omega}=0.16$. Point estimate based on reinfection studies [60–63] |
| $c_t$ – contact rate | 1, Equivalent to assuming contacts are back to baseline and stable |
| $v_i$ - Vaccine effectiveness against transmission (delta variant) | (0% to 35.0%)--dose 1,(0% to 69.0%)-dose 2 [15] |
| $v_a$ - Vaccine effectiveness against acquisition (delta variant) | (55% to 65%)-dose 1,(65% to 80%)-dose 2[14] |
| $v_d$ - Vaccine effectiveness against severe disease (delta variant) | (80% to 90%) dose 1,(95% to 99%)-dose 2 [14] |
| $v_\mu$ - Vaccine effectiveness against death (delta variant) | (90%-95%) -dose 1, (95% to 99%) -dose 2[14] |
| $1/r_{vp_i}$ - rate of vaccine progression to full efficacy | 14 days after each dose ($i = 1,2$) [64] |
| **Observation model parameters and data** |  |
| Number of people, by age group, who would test PCR positive on day $n$, $(P^+)_n$. | Dynamic |
| Number of people, by age group, who were observed to test PCR positive on day $n$, $(P_{obs}^+)_n$. | Data |
| Number of people, by age group, who would test as sero-converted on day $n$, $(S^+)_n$. | Dynamic |
| Number of people, by age group, who actually test as sero-converted on day $n$, $(S_{obs}^+)_n$. | Data |
| Probability that an infected individual would test PCR positive on day $t$ after infection, $Q_{PCR}(t)$ | $Q_{PCR}(t) = f_{onset} Q_\Gamma(\tau)$ where $Q_\Gamma(\tau)$ was the tail function of a gamma distribution fitted to data given in [65] and is the probability function of onset of symptoms post-infection [49]. |
| Probability that an infected individual would be detectably seropositive on day $t$ after infection, $Q_{sero}(t)$ | $Q_{sero}(t)$ is linearly increasing over 26 days to saturate at 92.7% sensitivity, based on report delay in seroconversion [65]and maximum sensitivity of serological assay [66]. |
| Relative bias in favour of selecting a PCR positive individual for testing, $\chi$ | Inferred from data |
| **Clinical outcome parameters** |  |
| The age-dependent probability of death given severe infection, $\mu_a^D$ | Inferred from data |
| The age-dependent probability of critical disease given severe infection, $\mu_a^C$ | Inferred from data |
| The age-dependent probability of severe disease given severe infection, $\mu_a^S$ | Inferred from data |
| Variant specific risk of death, $\psi_a^D$ | Inferred from data |
| Variant specific risk of critical disease, $\psi_a^C$ | Inferred from data |
| Variant specific risk of severe disease, $\psi_a^S$ | Inferred from data |

### Economic Evaluation

#### Equations Productivity losses

Productivity losses was calculated using the following equation:

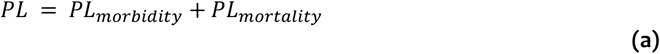

*Where*: PL=productivity losses; PL_morbidity_=productivity loss due to morbidity; PL_mortality_=productivity loss due to mortality

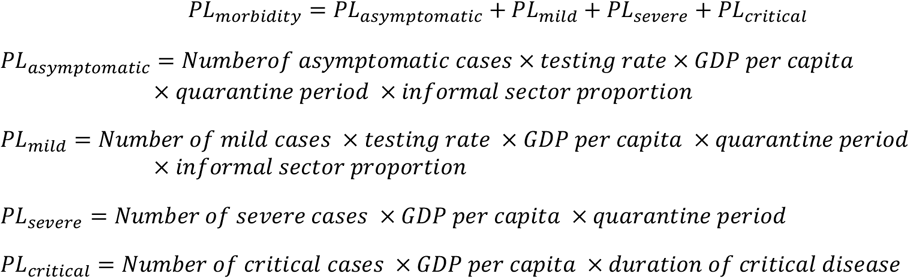

The testing rate was used to apportion the number of asymptomatic and mild cases who are tested. Further the proportion of informal sector is used to apply lost productivity on asymptomatic/mild cases that are in the informal sector, given the assumption that only those in informal sector are likely not to be productive as they isolate. Lastly, the duration of disease is used where the duration of illness is more than the 14 day quarantine period.

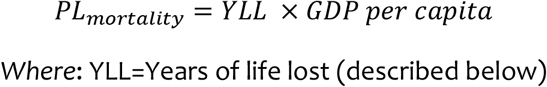

### Disability adjusted life years (DALYs)

Disability adjusted life years (DALYs) was calculated using the equation:

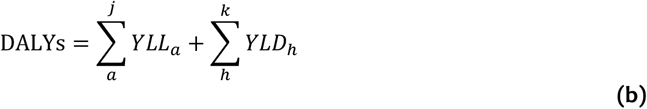

*Where*: *a* is the age at death; *j*= number of age groups; *h* are the health states; *k* =number of health states.

YLL is estimated as [22]:

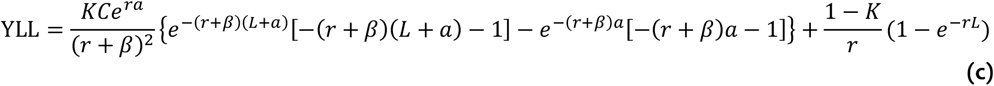

*Where*: K=age weighting modulating factor; C=adjustment constant for age weights; r=discount rate; a=age of death; β=parameter from the age weighting function; L=standard life expectancy at age of death

Years lost due to disability (YLD) is estimated as [22]:

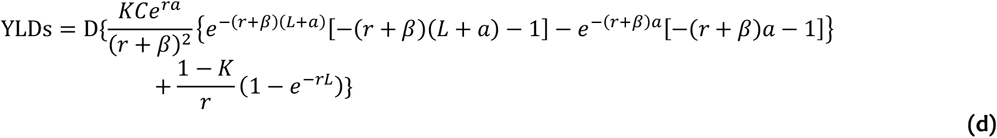

*Where*: K=age weighting modulating factor; C=adjustment constant for age weights; r=discount rate; a=age of onset of disability; β=parameter from the age weighting function; L=duration of disability; D=disability weight

**Table S2:** Statistical distributions for the probabilistic sensitivity analysis

|  | Statistical distribution* |
| --- | --- |
| **Costs** |  |
| Vaccine delivery cost per patient at 30% coverage | Gamma (10055,6.1E-4) |
| Vaccine delivery cost per patient at 50% coverage | Gamma (13679,3E-4) |
| Vaccine delivery cost per patient at 70% coverage | Gamma (9221,4E-4) |
| Unit treatment cost of an asymptomatic patient | Gamma (103.1,9.6E-4) |
| Unit treatment cost of a mild case | Gamma (105.1,1.8E-1) |
| Unit treatment cost of a severe case | Gamma (105.1,1.2) |
| Unit treatment cost of a critical case | Gamma (105.1,5.7) |
| **Health outcomes** |  |
| Disability weight for a critical case | Beta (102.8,54.2) |
| Disability weight for a severe case | Beta (21.6,140.8) |
| Disability weight for a mild case | Beta (21.3,396.2) |
| Length of hospitalization for severe episode | Gamma (53.3,3.6E-4) |
| Length of ICU stay for critical episode | Gamma (53.3,3.6E-4) |

\*The form of the distributions was chosen according to the nature of the parameter. (i.e. parameters describing the probability of health outcomes occurrence of an event are given by a beta distribution, while the values of the vaccine delivery costs and length of hospitalization/ICU stay of the various scenarios were described using a gamma distribution).

**Figure S6:**
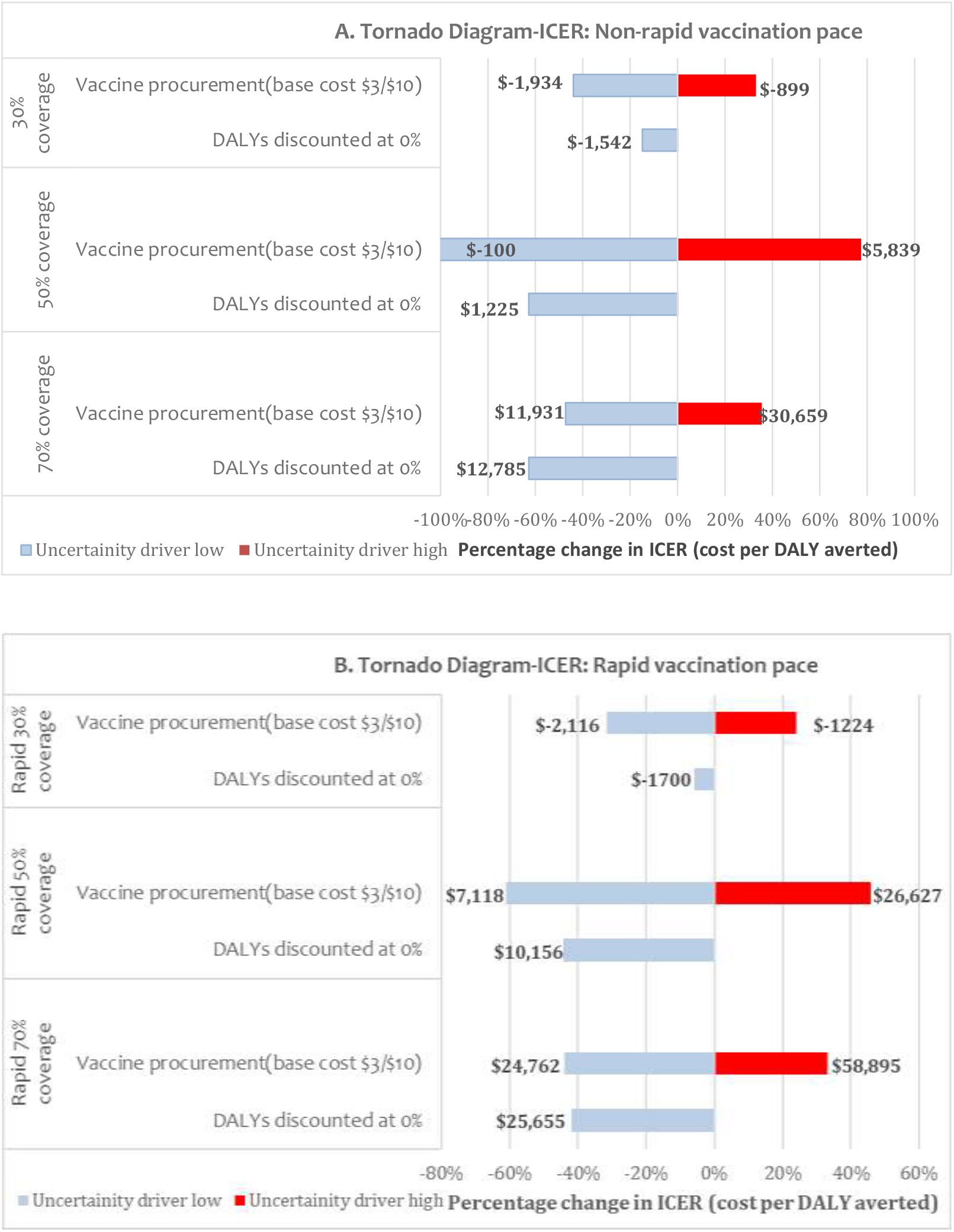
One-way sensitivity analysis of vaccine prices and discounting rate on ICERs

**Table S3:** Projected costs and the cost-effectiveness of different vaccination strategies in Kenya from a health system’s perspective

|  | Economic Outcomes |  |  |  |  |
| --- | --- | --- | --- | --- | --- |
| | Total vaccine costs<br>(\$ million)<br>Median (2.5<sup>th</sup> to 97.5<sup>th</sup><br>percentile) | Total treatment costs<br>(\$million)<br>Median (2.5<sup>th</sup> to 97.5<sup>th</sup><br>percentile) | Total health care costs<br>(\$million)<br>Median (2.5<sup>th</sup> to 97.5<sup>th</sup><br>percentile) | Total DALYs<br>(thousands)<br>Median(95%CI) | ICER, USD per<br>DALY averted<br>Mean (95%CI) |
| **Non-rapid vaccination pace (administered within 1.5years)** |  |  |  |  |  |
| No vaccination | - | 313<br>(269 to 405) | 313<br>(269 to 405) | 247<br>(243 to 252) | - |
| 30% coverage | 236<br>(233 to 238) | 157<br>(135 to 199) | 393<br>(371 to 436) | 114<br>(110 to 118) | 555<br>(553 to 557) |
| 50% coverage | 323<br>(321 to 324) | 140<br>(120 to 177) | 463<br>(444 to 501) | 101<br>(97 to 104) | 5,195<br>(5,190 to 5,199) |
| 70% coverage | 443<br>(441 to 445) | 133<br>(115 to 169) | 576<br>(560 to 612) | 96<br>(92 to 100) | 24,535<br>(24,514 to 24,557) |
| **Rapid vaccination pace (administered within 6 months)** |  |  |  |  |  |
| No vaccination | - | 313<br>(269 to 405) | 313<br>(269 to 405) | 247<br>(243 to 252) | - |
| 30% coverage | 236<br>(233to 238) | 129<br>(111 to 163) | 366<br>(347 to 399) | 93<br>(89 to 96) | 291<br>(290 to 295) |
| 50% coverage | 323<br>(321 to 324) | 124<br>(107 to 157 ) | 447<br>(430 to 481) | 88<br>(85 to 92) | 20,157<br>(20,126 to 20,128) |
| 70% coverage | 443<br>(441 to 445) | 121<br>(104 to 152) | 564<br>(549 to 596) | 86<br>(82 to 89) | 46,150<br>(46,025 to 46,274) |

Costs=rounded off to the nearest 1,000,000; Total DALY rounded off to the nearest 1,000; ICERs=rounded off to the nearest whole number

